# GWAS meta-analysis reveals dual neuronal and immunological etiology for pain susceptibility

**DOI:** 10.1101/2021.08.23.21262510

**Authors:** Evelina Mocci, Kathryn Ward, Susan G. Dorsey, Seth A. Ament

**Affiliations:** Department of Pain & Translational Symptom Science, University of Maryland School of Nursing, Baltimore MD; Center to Advance Chronic Pain Research (CACPR), University of Maryland Baltimore, Baltimore MD; Institute for Genome Sciences, University of Maryland School of Medicine, Baltimore MD; Department of Psychiatry, University of Maryland School of Medicine, Baltimore MD

## Abstract

Chronic pain is at epidemic proportions in the United States, represents a significant burden on our public health system and is coincident with a growing opioid crisis. While numerous genetic risk factors have been identified, its genetic basis remains poorly understood. Here, we conducted a meta-analysis of genome-wide association study (GWAS) summary statistics from seventeen pain susceptibility traits in the UK Biobank. This analysis revealed 99 genome-wide significant risk loci, of which 62 have not been previously associated with a pain-related trait. Risk loci were enriched for genes involved in neurological and inflammatory pathways. Two-sample Mendelian randomization indicated that depression, neuroticism, and immunological traits mediate many of these effects. These analyses double the number of known risk loci for pain susceptibility and support dual causation from neuronal and immunological genes, providing leads toward targets for novel pain medications.

## INTRODUCTION

Chronic pain is a public health epidemic, costing more than $600 billion in healthcare and lost work wages ^1^. Some of the most common chronic pain conditions include low back pain ^2^ and headache ^3^. Several lines of evidence suggest substantial individual differences in pain susceptibility, mediated in part by genetic factors. Pain patients with seemingly identical injuries often report different pain intensities, both in clinical settings and in experimental studies where pain stimuli are strictly controlled ^4–6^. Individual differences in pain response and susceptibility to develop chronic pain conditions are at least in part inherited ^7,8^. Twin studies focused on several of the most common pain-related phenotypes estimated broad-sense heritability of 39-58% for migraine ^9–11^, 32-50% for back pain ^12,13^, and 21% for sciatica ^14^. A more recent study on pain heritability, analyzed nearly 7,600 samples from 2,500 families controlling for shared environmental factors, found broad-sense heritability of 16% for chronic pain, and the estimate increased to 30% when the analysis was restricted to severe chronic pain.^15^

The study of genetic differences in pain intensity and susceptibility to develop chronic pain conditions has been hampered, however, by inadequate sample sizes and a lack of consistent findings. With the availability of the UK Biobank (UKBB) data, there is now a large study population available (∼500,000), with extensive phenotype data, to conduct well-powered genetic association studies of pain ^16^. Recent studies have leveraged the UKBB database to conduct pain GWAS of headache (N=223,773) ^17^, osteoarthritis (N=455,221) ^18^, low back pain (N=487,409) ^19^ and (N=158,000) ^20^, knee pain (N=171,516) ^21^, neuropathic pain (N=150,000) ^22^ and multisite chronic pain (N=380,000) ^23^. Each study identified known and novel loci associated with the specific pain condition under study, but few overlapping loci were identified across pain conditions. This is surprising, given that similar pain phenotypes should share at least some genetic correlation^24^. If so, a joint analysis of multiple pain-related traits could increase statistical power and identify novel loci. Here, we present a joint analysis of seventeen pain-related phenotypes from UKBB, including short-term and chronic pain, generalized and site-specific traits and proxies such as pain relief medications.

## RESULTS

### Discovery of 99 risk loci for pain susceptibility in the UK Biobank

To gain insight into the genetic architecture of pain phenotypes, we started by estimating the genetic correlations (rg) among 17 pain-related UKBB traits with substantial heritability (**Fig. 1A; Supplementary Tables 1 and 2**); genetic correlations were strong among pain reported in different sites of the body, including knee, hip, leg, back, and neck-shoulder, as well as between pain at individual sites and the aggregate trait, pain anywhere in the body (rg ranging from 0.81 to 0.93). (**Fig. 1B, Supplementary Table 3**). Headache was correlated with categories of pain medications, including paracetamol and ibuprofen, as well as with the use of any pain medication. Chest pain and use of aspirin were genetically correlated with each other but not with other traits, most likely reflecting the use of aspirin as a blood-thinner and reflecting distinct genetic causes for pain associated with cardiovascular diseases. Taken together, these analyses suggest a shared genetic component for susceptibility to many, but not all, forms of pain.

**Figure 1.**
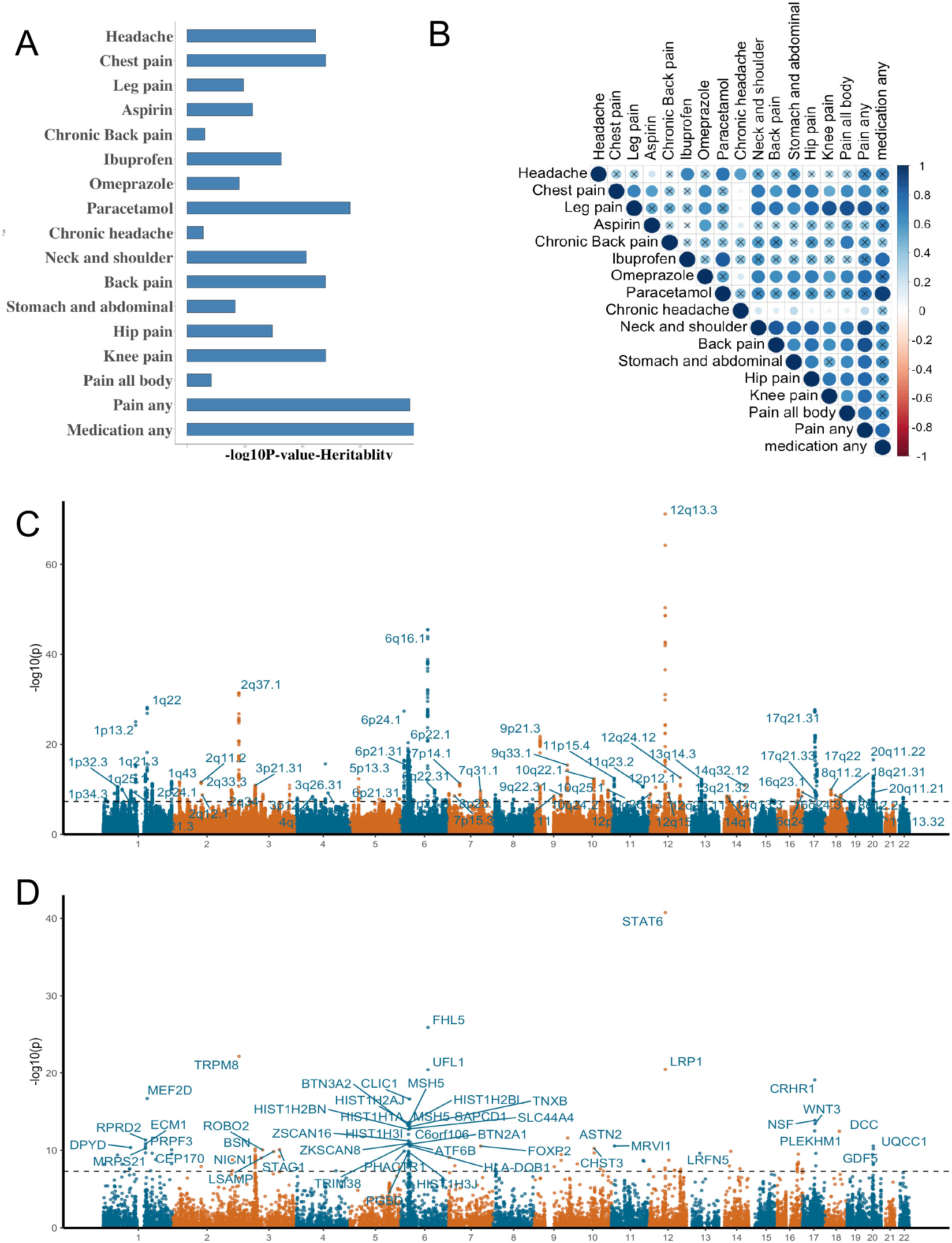
Joint GWAS analysis of 17 pain-related traits. **A**. SNP heritability estimates for 17 pain-related traits in the UK Biobank. Y axis indicates the trait and the X-axis indicates the -log_10_(p-value) of the heritability estimate as calculated with LD Score Regression. **B**. Pairwise genetic correlation estimates between all 17 pain related traits. red = negative genetic correlation; blue = positive genetic correlation. The size of the circles is proportional to the value of genetic correlation, and non-significant correlations are indicated by an “X”. **C**. Manhattan plot derived from joint analysis of the 17 traits with PLEIO. The x-axis indicates chromosomal position and the y-axis indicates -log_10_(p-value). **D**. Manhattan plot for gene-based analysis of joint GWAS summary statistics, computed with MAGMA.

Joint analysis of genetically correlated traits improves statistical power to identify risk loci. We therefore conducted a joint GWAS analysis of the 17 pain related traits using PLEIO ^25^ and identified 1,770 genome-wide significant pain-associated SNPs (*P* < 5e-8) at 99 approximately linkage-disequilibrium (LD) independent loci (**Fig. 1C, Supplementary Table 4**). The quantile-quantile plot indicated substantial deviation from a null distribution (**Supplementary Figure 1**). LD score regression (LDSC) ^26^ indicated that this is primarily due to polygenicity rather than to confounding factors (LDSC intercept = 0.84; ratio = 0). Overall, ∼13% of trait variance (h2) was explained by the genotyped and imputed SNPs. This represents a substantial increase in power compared to GWAS of individual pain-related traits. At 51 of the risk loci in the joint analysis, none of the underlying GWAS of individual traits reached genome-wide significance (**Supplementary Table 5**). At an additional 37 loci, GWAS of only one of the 17 traits reached significance, primarily the risk loci for headache, use of any pain medication, use of paracetamol, or pain at any site in the body. Only 11 loci reached genome-wide significance for more than one trait in the original GWAS, prior to joint analysis.

Sixty-two of the 99 risk loci have not previously been described in published GWAS of pain-related traits (**Supplementary Table 6**). However, 37 of our 99 loci overlap with previously reported risk loci for pain phenotypes, most of which arose from single trait analysis of the UKBB sample (**Supplementary Table 7**). Of these, 20 loci overlapped a previously reported risk locus for migraine headache ^3,27–32^, while 10 loci were previously reported for multisite chronic pain ^23^. Three loci (1q21.3, 9q33.1 and 20p11.23) were found to be associated with both migraine and multisite chronic pain ^23,27–31^, while 17q22 was formerly described as associated with neck or shoulder pain ^33^ and multisite chronic pain GWASs ^23^. Chromosome 7q31.1 was previously associated with neck pain or shoulder pain, 12p12.1 with chronic back pain ^20^, and 20q11.22 with knee pain ^21^. In summary, the joint analysis method more than doubled the number of risk loci that were able to be identified using single pain-related traits.

Published GWAS of pain susceptibility performed outside the UKBB sample were orders of magnitude smaller, precluding a classical replication analysis. In light of this, we undertook pseudo-replication by applying joint GWAS analyses to males and females in the UKBB cohort separately, and we compared these sex-specific results to our primary analysis. The female-specific analysis identified 25 genome-wide significant risk loci, with no inflation of test statistics (Lambda GC: 1.0285, LDSC intercept: 0.9971) (**Supplementary Figure 2A, Supplementary Figure 3A)**. All female-specific loci were also risk loci in the primary analysis. Ninety-eight of the 99 loci from the primary analysis were replicated in the female-specific analysis at a nominal p-value < 0.05 (**Supplementary Table 8)**. Similarly, the male-specific analysis revealed 16 genome-wide significant risk loci, with no inflation of test statistics (Lambda GC: 1.0225, LDSC Intercept: 1.0006) (**Supplementary Figure 2A, Supplementary Figure 3B)**. All male-specific loci were also risk loci in the primary analysis. Ninety-five out of 99 loci from our primary analysis were replicated at *P* < 0.05 (**Supplementary Table 8**). These results suggest that most risk loci for pain are reproducible in both male and female subjects.

### Pain susceptibility risk loci are enriched for genes with neurological and immunological functions

We performed gene-based analyses with MAGMA^34^ to identify functional categories of genes enriched at risk loci for pain. MAGMA analysis of summary statistics from our joint analysis identified 300 pain-associated genes at a genome-wide significance threshold, *P* < 2.8×10-6, adjusting for 17,620 protein coding genes (**Figure 1D, Supplementary Table 9**). Of these, 46 were the closest gene to an LD-independent lead SNP at one of the 99 genome-wide significant risk loci, while the remaining genes were distal to these SNPs or located outside the genome-wide significant risk loci (**Supplementary Table 10)**.

First, we hypothesized that genes associated with risk for pain would be enriched for genes that have previously been shown to influence pain sensitivity. To test this hypothesis, we considered 353 human pain genes discovered in clinical studies, candidate gene, and GWAS analyses 35, as well as 434 pain genes identified in mouse studies 36. MAGMA gene set enrichment analysis applied to human pain genes revealed a statistically significant enrichment of these known pain genes at our identified risk loci (*P* = 1.65e-4) **(Supplementary Table 11)**. Ninety genes included in the human pain genes dataset reached nominal significance in our MAGMA analysis (p < 0.05). These genes have most frequently been reported to be associated with migraine and analgesia related phenotypes, as well as with nociception, neuraxial pain, and temporomandibular disorder. We also found a significant enrichment for the mouse knockout pain genes (*P* = 2.0e-2). Notable genes with evidence from functional studies include ion channels (e.g., *TRPM8, P* = 6.4e-23), ligand-gated receptors (e.g., *LRP1, P* = 3.1e-21), immune system (e.g., *H2AB1, P* = 2.96e-11) and extracellular matrix (e.g., *NCAM1, P* = 2.0e-9) (**Supplementary Table 11**). These analyses support the relevance of the risk loci identified in this study to known genes and pathways associated with pain sensitivity.

However, many of our identified risk loci do not contain a known pain gene, suggesting that they reflect novel mechanisms. Thus, we next hypothesized that genes associated with an increased risk for pain may be enriched for expression in specific tissues and cell types. Non-pain risk loci were significantly overrepresented (FDR < 0.05) near genes expressed in several brain regions including the cerebellum, frontal cortex, anterior cingulate cortex, and nucleus accumbens (**Supplementary Figure 4)**. These results suggest that heritable factors contributing to pain susceptibility may act via mechanisms within the brain.

Finally, we performed unbiased gene set enrichment analysis with functional categories from Gene Ontology (GO)^37^, the Kyoto Encyclopedia of Genes and Genomes (KEGG)^38^, and Reactome ^39^ (**Supplementary table 12)**. These analyses revealed the strongest enrichments for gene sets related to neuronal development and function (e.g., neurogenesis, *P* = 3.2e-8, neuron differentiation, *P* = 1.4e-6, post synapse, *P* = 4.6e-7). Top pain-associated neurogenesis genes included *LRP1* (*P* = 3.1e-21), *UFL1M* (*P* = 3.7e-14), *WNT3* (*P* = 1.7e-13), *SLC44A 4* (*P* = 3.5e-13), *DCC* (*P* = 6.4e-11) and *GDF5* (*P* = 1.2e-10) (**Supplementary Table 13**). Additionally, we found significant enrichment in categories related to immunological functions.: e.g., chemokine secretion (*P* = 1.4-4), T-helper T-cell differentiation (*P* = 1.3-4) and reactome T-cell receptor (TCR) signaling, P= 1e-04); top pain-associated genes with immunological functions included *STAT6 (P=1*.*7e-41)* and *CRHR1 (P=8*.*4E-20)*. These results support dual neurological and immunological etiology for pain susceptibility.

### Genetic risk for pain is mediated by effects of SNPs on neuropsychiatric, immunological, and cardiometabolic traits

To further investigate the neurological and immunological roots of pain sensitivity, we considered the hypothesis that SNPs associated with pain have pleiotropic effects within those domains. First, we asked whether individual SNPs associated with pain susceptibility are also associated with risk for other phenotypes. We scanned pleiotropic associations of the lead SNPs and SNPs in LD with them (r2 > 0.6) among the SNPs associated with >4,000 non pain-related traits collected in the GWAS Atlas ^40^. We estimated that our SNPs were significantly overrepresented in the GWAS Atlas dataset (Fisher’s exact test: odds ratio = 69.9; *P* < 2.2e-16), providing initial evidence that our loci have pleiotropic effects on other phenotypes besides pain.

Interestingly, risk-associated SNPs at 19 of the 99 pain loci were pleiotropically associated with risk for a psychiatric or neurological trait (**Fig. 2A; Supplementary Table 14)**. The largest number of loci had pleiotropic associations with risk for major depression or neuroticism. Pleiotropic associations were also found for Parkinson’s disease and brain structure. This analysis also revealed pleiotropy with immunological traits, as 8 pain risk loci were reported associated with hematological traits related to erythrocytes, leukocytes, and platelets. We also found pleiotropic associations with cardiovascular and metabolic traits. However, examination of summary statistics from individual pain-related traits suggested that these latter associations are driven by SNPs that are associated with chest pain and use of aspirin, and thus may represent a distinct category from that of SNPs that are associated with other forms of pain. Therefore, this analysis primarily supports pleiotropic associations of pain with brain-related and immunological traits.

**Figure 2.**
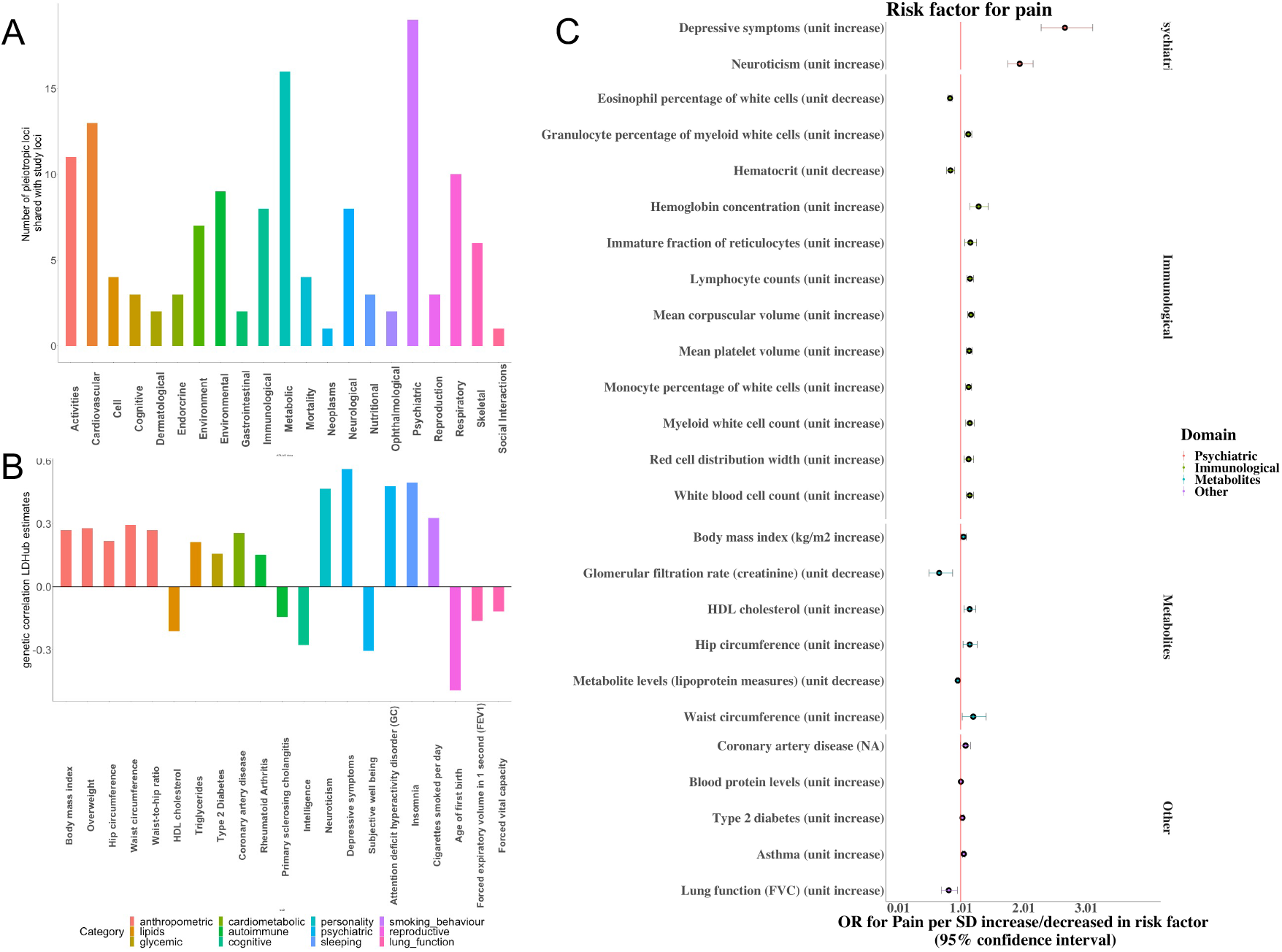
Pleiotropy, genetic correlation, and predicted causal interactions between pain susceptibility and non-pain traits. **A**. The number of pleiotropic loci shared between pain and non-pain traits collected in the GWAS Atlas database. **B**. Genetic correlations between pain and non-pain traits. Traits are grouped by domains indicated by bar color. **C**. Two-sample Mendelian Randomization results. The X axis indicates the odds ratio (OR) for pain and its 95% confidence interval (CI). The OR is computed per standard deviation (SD) increase or decrease of the risk factor. All risk factors for pain that have a significant causal relation to pain are listed in the Y axis, and they are grouped by domain.

We extended this analysis by computing genome-wide genetic correlations between pain susceptibility and neuropsychiatric, immunological, and metabolic traits with LDSC ^26^. We observed strong genetic correlation with personality traits such as neuroticism (genetic correlation range: 0.39-0.46) and several psychiatric traits, particularly those related to depression (genetic correlation range: 0.35-0.56) and attention deficit hyperactivity disorder (genetic correlation range: 0.30-0.48), insomnia (genetic correlation range: 0.45-0.50) and education-related traits (genetic correlation range: 0.30-0.45) (**Fig. 2B, Supplementary Table 15**). Likewise, several anthropometric traits related to BMI, metabolic traits based on glycemia, lipid and other metabolites were shown to be significantly correlated to our loci. Genetic correlations between pain and immunological traits were not as strong in this analysis. However, for several traits, including mean corpuscular volume (MCV) and mature and immature red cell traits, we detected nominally significant correlations (*P* < 0.05).

Next, we tested for possible causal relationships between pain and non-pain traits, focusing on traits for which we had identified either a pleiotropic association with a genome-wide significant SNP or significant genetic correlations. Two-sample Mendelian randomization ^41^ identified causal relationships primarily in five categories: psychiatric, immunologic, metabolites, environmental, and other (**Fig. 2C; Supplementary Figures 5-10; Supplementary Table 16)**. First, we found a significant causal effect of neuroticism and depression on the risk of experiencing pain. Second, we found bidirectional mediating effects between genetic factors contributing to increased blood concentrations of eosinophils, granulocytes, reticulocytes, monocytes, neutrophils, and platelets and risk for pain. Third, we observed that metabolic traits including both those linked to overweight/obesity such as high body mass index, elevated waist and hip circumference, and those linked to lipoprotein and other metabolites levels, were significantly associated to increased pain. Fourth, respiratory traits, such as asthma and lung function, which showed significant genetic correlation with our meta-analysis (**Fig. 2B)**, were found causally linked to pain; specifically, asthma caused an increase in pain, whereas elevated forced vital capacity (FVC) levels diminished pain. Fifth, among the other traits, we detected a significant causal link with coronary artery disease and type 2 diabetes, both of which were associated with a small but significant increase of pain susceptibility. For all traits, we confirmed that the classic inverse variance weighted method was supported by sensitivity tests. Additionally, by applying Cochrane Q heterogeneity and pleiotropy tests we were able to rule out the effect of risk factors, other than those analyzed, through which the genetic variant/s of interest impact on pain. In summary, we found that genetic risk for pain is mediated by effects of diverse non-pain traits, with many of the strongest effects from neuropsychiatric and immunological traits.

### Fine-mapping of pain-associated SNPs and their pleiotropic effects on genetically correlated traits

Next, we investigated these causal mechanisms and mediating effects of neuropsychiatric, immunological, and cardiometabolic traits at specific risk loci. We predicted the genes impacted by the SNPs at each risk locus based on proximity to risk-associated SNPs, as well as by integration with expression quantitative trait loci (eQTLs) and chromatin interaction data from the human cortex ^42^, finding a total of 664 genes at the 99 risk loci (**Supplementary Table 17, Supplementary Figure 11**). We fine-mapped potential causal SNPs and mediating effects of other traits using a combination of two-sample Mendelian randomization and functional annotation. These analyses identified 41 significant effects (**Supplementary Table 18, Fig. 3A**). Below, we highlight examples from eight of the 99 risk loci.

**Figure 3.**
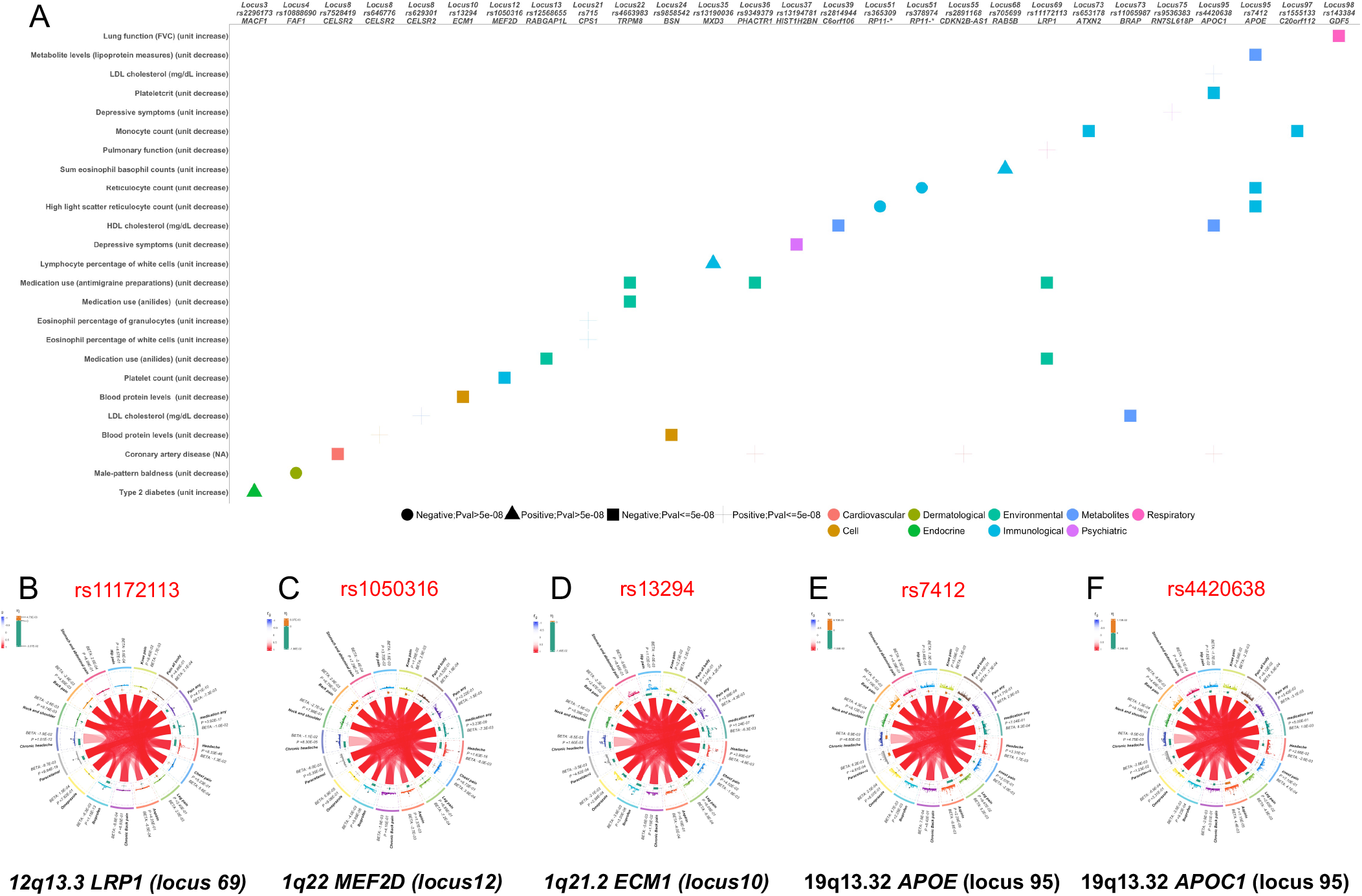
Annotation and pleiotropy of selected risk loci. **A**. Mediating effects of non-pain traits on risk for pain, derived from single SNP Mendelian randomization analysis. X-axis indicates genomic locus, SNP, and nearest gene. Non-pain exposure traits are indicated on the y-axis. Symbols and colors indicate direction of the effect and significance. **B-F**. Strength of associations to the 17 individual pain-related traits for four of the 27 loci represented in (A), Each plot provides information about each of the 17 traits at a specific locus. At center, blue and red arcs indicate the genetic correlation between the 17 pain-related traits. The innermost ring indicates the effect size and its direction of the leading locus SNP on pain for each one of the 17 traits. Yellow = increased risk of pain; green = protective effect. The size of the effect is proportional to the width of the line. The middle ring shows a region plot for the locus indicating the -log10(p-values) for associations of SNPs in this region with each trait. The darker point indicates the P-value of the lead SNP. Finally, in the outermost ring are reported the name of the trait, beta coefficient and standard error of the SNP effect, and p-value for genetic association.

The SNP most strongly associated with pain genome-wide was rs11172113 (*P* = 6.9e-72**; Fig. 3B)**. The alternate allele was associated with increased risk for pain and was previously shown to be associated with decreased expression of *LRP1* ^43^. Rs11172113 is also associated with an increased intron-excision splicing of an adjacent gene, *STAT6*. MR analysis suggests that the increase in pain associated with rs11172113 may be mediated by decreased pulmonary function. *LRP1* encodes the low-density lipoprotein receptor-related protein 1, while *STAT6* encodes for a transcription factor. Both *LRP1* and *STAT6* have well-known functions in inflammation, including in pulmonary function ^44,45^. Therefore, we interpret that this risk locus may impact pain susceptibility by modulating inflammatory processes.

The rs1050316 alternate allele, located in the 3’ UTR of *MEF2D*. was associated with a substantial decrease in risk for pain (OR=0.34, 95%CI=0.28-041, *P* = 6.2e-29); **Fig. 3C)** and previously with decreased expression of *MEF2D* ^43^. MR analysis suggested that this effect may be mediated by a decreased platelet count and decreased platelet volume. MEF2D is a transcription factor that is highly expressed in macrophages and other myeloid cells, with important roles in the regulation of innate immune states 46, further supporting that this locus impacts the function of cells in the immune system. rs13294 (*P* = 1.4e-13; **Fig. 3D**) encodes a missense variant in *ECM1* (Extracellular Matrix Protein 1, ECM1 Gly442Ser) in LD with an independently significant SNP at 1q21.2. The rs13294 alternate allele was associated with decreased risk of pain; MR analysis suggests that this effect may be mediated by a decrease in blood protein levels, again implicating effects on immune cells. *ECM1* is highly expressed in macrophages and is involved in the regulation of inflammatory states in diseases such an inflammatory bowel disease, as well as cancer ^47^. We previously identified extracellular matrix (ECM) as the most significant biological pathway associated to genes found differentially expressed in a transcriptome-wide study comparing chronic low back pain patients with healthy controls ^48^. SNPs located within genes included in this pathway are also known to be associated with human back pain and across both mouse and human. ECM organization is a central mechanism associated with chronic pain ^49^.

Single SNP MR analyses also highlighted a locus on chromosome 19q13.32 with two independent SNPs, rs7412, a missense variant in APOE (**Fig. 3E**) and rs4420638 (**Fig. 3F)** located downstream APOC1. This locus demonstrated high pleiotropy in our study; the association between pain and these two different SNPs was mediated by effects shown in several immunological, metabolic, and cardiovascular traits (**Supplementary table 18, Fig. 3A**). APOE isoforms are well-known risk factors for Alzheimer’s disease. Most carriers of the ancestral rs7412 T allele express the APOE ε2 isoform, while the rs7412 C allele, in combination with the C allele of a second SNP, rs429358, define the Alzheimer’s risk-associated ε4 isoform^50^.

Finally, our analysis detected genomic regions that mediate the effect of psychiatric factors on pain. Two pain-susceptibility loci discovered in this study were linked to depressive symptoms. On chromosome 6p22.1, within the extended major histocompatibility (MHC) region, the minor allele G of SNP rs13194781 was associated with decreased depressive symptoms, as well as with a protective effect for pain. Although extended LD in this region confounds fine-mapping, rs13194781 is an eQTL with effects on several nearby genes. A region on chromosome 13q14.3 showed association with increased depressive symptoms and increased risk of developing pain.

## DISCUSSION

A meta-analytic approach enabled us to identify 62 novel genomic regions associated with pain and to confirm 37 of those already reported in the literature. Gene set enrichment analysis, genetic correlation analysis, and genetic mediation analyses provided biological insights into underlying mechanisms. Based on these results, we postulate that many of the genetic factors contributing to pain susceptibility fall into by two broad categories: neurological effects that may alter the perception of pain within the brain and immunological effects that may increase inflammatory tone.

Our analyses establish neuronal genes and neuropsychiatric mediation effects in genetic risk for pain. Gene set enrichment analyses revealed enrichments for genes expressed in several brain regions and for functional categories related to the development and functions of neurons. Lead SNPs at 19 of the 99 pain risk loci identified in the present study had pleiotropic effects on psychiatric phenotypes, particularly depression, and we identified a positive genetic correlation between pain susceptibility and depression. Mendelian randomization suggested that depression and other mood-related traits mediate genetic risk for pain. These observations extend previous observations regarding genetic risk for pain-related phenotypes, notably multisite chronic pain ^23^, identifying additional shared loci. Somatization, defined as the tendency of some individuals to experience and communicate somatic distress in response to psychosocial stress, occurs frequently in patients with major depression and related disorders ^51^, perhaps explaining depression’s genetic mediation of pain susceptibility. Chronic pain is also associated with changes in brain structure and function independent of depression, detectable through human brain imaging ^52^.

Genes at pain risk loci implicate mechanisms in both the central and peripheral nervous system. For instance, we found a significant association at *TRPM8*, encoding a cold-sensitive cation channel expressed primarily in peripheral nerves. Knockdown of *Trpm8* in mice demonstrated an essential role in the suppression of sensitized pain responses ^53^. By contrast, a second neuronally expressed risk gene, *NCAM1*, encoding neuronal cell adhesion molecule 1, is expressed primarily in the central nervous system and is involved in cell adhesion and synaptogenesis. Cortically expressed NCAM1 has been shown to regulate synaptic reorganization after peripheral nerve injury ^54^. Thus, our data implicate multiple neurological mechanisms in pain susceptibility

Likewise, inflammatory and immunological mechanisms were supported by multiple analyses. Gene set enrichment analyses revealed an over-representation of risk loci near genes with functions in immune cells. We detected pleiotropic effects on pain and immunological traits at eight loci. Mendelian randomization supported causal mediating effects for diverse traits that are likely to reflect changes in inflammatory tone. Chronic activation of immune cells in damaged tissues (i.e., inflammation) is known to cause hypersensitivity of peripheral pain-sensing neurons by several mechanisms ^55^. This inflammatory pain normally resolves as tissues heal, while a failure of inflammation to resolve in conditions such as arthritis contributes to chronic pain ^55^. However, it is notable that genetic risk factors for pain were not genetically correlated with risk for chronic inflammatory disorders such as Crohn’s disease or rheumatoid arthritis. This result is consistent with previous findings ^23^ and suggests that distinct aspects of inflammatory tone contribute to chronic pain. Instead, we detected mediating effects of diverse hematological traits, as well as with traits associated with persistent systemic inflammation such as deficits in pulmonary function. Several genes at risk loci have known roles in modulating inflammatory pain. In addition to examples described above, *CRHR1*, encoding corticotropin-releasing hormone (CRH) receptor 1 contributes both peripherally to the excitation of pain-sensing neurons and centrally in setting inflammatory tone within the hypothalamus-pituitary-adrenal axis ^56^.

There are limitations associated with our approach; namely that our choice of phenotypic coding could influence interpretation. First, a small number of individuals who did not respond to pain-related questions are included in the analysis, coded as “population controls”. The inclusion of population controls maximizes sample size. In practice, since response rates to all the pain-related traits were very high, this affects at most a few percent of individuals in the analysis. Second, individuals with chronic pain for 3+ months are compared to individuals who experienced pain for shorter amounts of time (i.e., “pain-exposed” controls). We believe this definition leads to a cleaner phenotype than a comparison with individuals that did not experience pain at all, but we note that it leads to a small sample size for the chronic pain traits. Third, the analysis treats each trait separately. As such, certain individuals are treated as controls for one trait and cases for another; i.e., an individual who only experiences pain in their back but not in their chest would be considered a case for back pain but a control for chest pain. While potentially confusing, we feel that this is a justifiable choice to explore potential differences between pain location risk susceptibility. More broadly, assessing pain in biobank-scale cohorts inevitably leads to a tradeoff in terms of the depth of phenotyping, and there will be value in continuing to develop cohorts with more detailed clinical assessments of pain.

Joint analysis of multiple pain-related traits led to a substantial boost in statistical power, more than doubling the number of known risk loci for pain. By definition, the pain phenotype that we are studying is very broad. The increased power suggests that genetic risk factors are shared among many pain-related traits. We can infer, therefore, that the neurological and immunological signals we detect predispose to pain very generally. Presumably, there are additional risk factors that contribute to more specific forms of pain. Increasing sample sizes through incorporation of data from additional cohorts will likely lead to the identification of additional loci.

The management of chronic pain represents a substantial medical challenge. Many existing pain medications are highly addictive, especially opioids. Thus, there is an urgent need to develop new, non-addictive pain medications. Our discovery of scores of pain risk loci has the potential to identify novel therapeutic targets. Encouragingly, genes at several of the risk loci are already being targeted therapeutically (e.g., TRPM8). Genes at many other loci are not yet being targeted but could represent novel strategies. To exploit this potential, it will be essential to continue to refine our understanding of the underlying mechanisms through fine-mapping and functional experiments.

## METHODS

### Sample and phenotype selection

Summary statistics were available from GWAS of >4,200 traits in 361,194 unrelated samples (194,174 females and 167,020 males) of ‘white British’ ancestry from the UKBB cohort ^57^ (http://www.nealelab.is/uk-biobank), including >13 million genotyped and imputed SNPs. Pain-related phenotypes were selected using the keyword ‘pain’. We selected traits with sample size > 10,000 and statistically significant heritability estimates, z>4, based on LD Score Regression (LDSC) analyses performed by the Neale lab (https://nealelab.github.io/UKBB_ldsc/h2_browser.html) 58. We calculated pairwise genetic correlations among the selected traits using LDSC 58 and set the value of 0.35 as threshold to remove traits with low genetic correlation. However, all identified pain-related traits showed pairwise rg >= 0.35 with at least one other trait, resulting in a final set of 17 pain-related traits (details in **Supplementary Table 1)**. GWAS summary statistics for all traits were downloaded from http://www.nealelab.is/uk-biobank, GWAS round 2. Technical details on samples, quality control, and statistical modeling are described at https://github.com/Nealelab/UK_Biobank_GWAS. In the current study, we considered only autosomal markers with minor allele frequency (MAF) >=0.01, without significant deviation from Hardy Weinberg Equilibrium (p-value>0.00001).

### Meta-analysis of UKBB 17 pain related GWASs

Combined analysis of the 17 selected pain related traits was carried out using PLEIO 25 (https://github.com/hanlab-SNU/PLEIO). PLEIO corrects for environmental correlations due to sample overlap. Here, we estimated environmental correlation as the intercept estimate of the genetic covariance computed by applying cross-trait LD Score regression ^58^. In addition, PLEIO relies on standardized trait effect sizes to express all phenotypes in the same units. Since our phenotypes are binary, we standardized effect sizes by converting them to a liability scale, assuming prevalence in the UKBB cohort is equivalent to that of the broader UK population. We first ran the meta-analysis in the whole sample and then, to evaluate robustness of findings and detect sex-specific effects, we also analyzed GWAS summary statistics from males and females, separately. We used the joint analysis statistics to estimate pain heritability explained by the additive effects of the SNPs using LD score regression^26^.

### Risk loci and gene mapping

Post-GWAS analyses of risk loci, gene mapping, and functional annotation were performed with FUMA SNP2GENE ^59^. Risk loci were defined around genome-wide significant SNPs (p-value < 5e-8) using the following parameters: r^2^ threshold to define independent significant SNPs >= 0.6; 2nd r^2^ threshold to define lead SNPs ≥ 0.1; maximum distance between LD blocks to merge into a locus < 250 kb. LD was based on the 1000 Genomes Phase 3 reference panel population. Additional SNPs at each locus were defined as those with a p-value < 0.05 and r^2^ ≥ 0.1 with an independent significant SNP. SNPs in LD with an independent significant SNP were functionally annotated using ANNOVAR ^60^, based on Ensembl build 85. Gene mapping was performed using three complementary approaches. Positional mapping selected any gene +/- 10kb from the tag SNP. Expression quantitative trait locus (eQTL)-based mapping used the PsychENCODE database ^42^. Genes were selected if their expression was significantly (FDR<0.05) associated with the tag SNP allele and expression was higher than 0.1 FPKM (Fragments Per Kilobase of transcript per Million mapped reads) in at least 10 samples ^59^. In this analysis we considered only cis-eQTLs, defined as genes located < 1MB from the tag SNP. For top risk loci, we also manually annotated multi-tissue eQTLs from the Genotype-Tissue Expression Consortium ^43^. Chromatin interaction mapping used Hi-C data from PsychENCODE ^42^ and considered only one-way enhancer-promoter interactions (FDR < 0.05) within 500 kb of the tag SNP.

### Gene set enrichment analysis

We performed gene set enrichment analysis of GWAS summary statistics using MAGMA ^34^, which is implemented within FUMA, as well as a standalone package. Initial steps, including SNP annotation and calculation of gene-based p-values were performed in FUMA using default parameters and 1000 Genomes Phase 3 as reference. Next, competitive gene-set analysis was carried out with gene sets annotated using MSigDB v7.0 (https://www.gsea-msigdb.org/gsea/msigdb ^61–63^. Similarly, a tissue expression analysis was run where the previously computed gene-based p-values were conditioned by the average expression of genes per tissue type; FUMA implements this analysis using both 30 general tissue types and 53 specific tissue types ^59^ from the GTEx consortium. In addition, we tested if the genes detected in our study were enriched for genes previously found linked to pain. For this purpose, we used two lists of genes: (i) genes derived from human pain genetics ^35^; (ii) pain genes identified in murine knockout studies ^36^.

### Identification of pleiotropic effects for individual SNPs

We looked up pleiotropic associations to non-pain-related traits in the GWAS Atlas ^40^ for each individual significant SNP associated with pain, as well as other pain-associated SNPs in LD with an independent significant SNP. The GWAS Atlas database catalogs genome-wide significant SNPs from 4,756 GWAS across 3,302 unique traits, which are categorized into 28 domains.

### Genetic correlation

LDSC was used to calculate genetic correlations between pain and other traits. We conducted an unbiased analysis considering all traits in the LDHub database with the exception of those from the UKBB cohort ^64^, as well as selected immunological traits for which we identified pleiotropic effects in the GWAS Atlas ^40^.

### Two-sample Mendelian Randomization (MR)

Two-sample Mendelian randomization was performed from GWAS summary statistics of traits for which we found at least one pleiotropic effect in the GWAS Atlas using the TwoSampleMR R package ^41^. This method uses multiple variants, weighted by their effect and combined into a multi-locus allele score ^41^.

TwoSampleMR implements several sensitivity tests, including MR Egger, weighted median, and mode Mendelian randomization in addition to the primary inverse variance weighted (IVW) test, which assumes no horizontal pleiotropy ^65^. Each pair of traits was tested, considering pain as exposure and traits as outcome, and vice versa. We considered MR results as plausible when the IVW test and the majority of the sensitivity tests were significant or trended toward significance. Different directionality of statistical tests, even if significant, was not considered a meaningful result. When the results suggest pleiotropy, both IVW and MR Egger were tested for heterogeneity among their single SNPs estimates using Cochran Q heterogeneity test ^66^. Pleiotropy was measured from the intercept term in the MR Egger method and interpreted as an estimate of the average pleiotropic effect across all genetic variants 65; an intercept term that differs from zero is indicative of overall directional pleiotropy. In addition, to detect the effect of each of the SNPs included in the multi-locus allele on pain, we performed twoSampleMR analysis on each SNP individually using the Wald ratio method ^41^. We considered results to be significant only when the SNPs reached genome-wide significance for association both with pain and with the predicted mediating factor.

### Data Availability

Complete GWAS summary statistics will be made available through the GWAS Catalog (https://www.ebi.ac.uk/gwas/). All other data and results are contained in the manuscript and supplementary files.

## ACKNOWLEDGEMENTS

This work was supported by a grant from the National Institute of Nursing Research to S.G.D. (P30 NR016579).

