## Supplementary Figures 1-11 for "GWAS meta-analysis reveals dual neuronal and immunological etiology for pain susceptibility"

**SUPPLEMENTARY INFORMATION**

This supplement includes 11 supplementary figures and 18 supplementary tables presented as Excel files.

**Supplementary Figures**

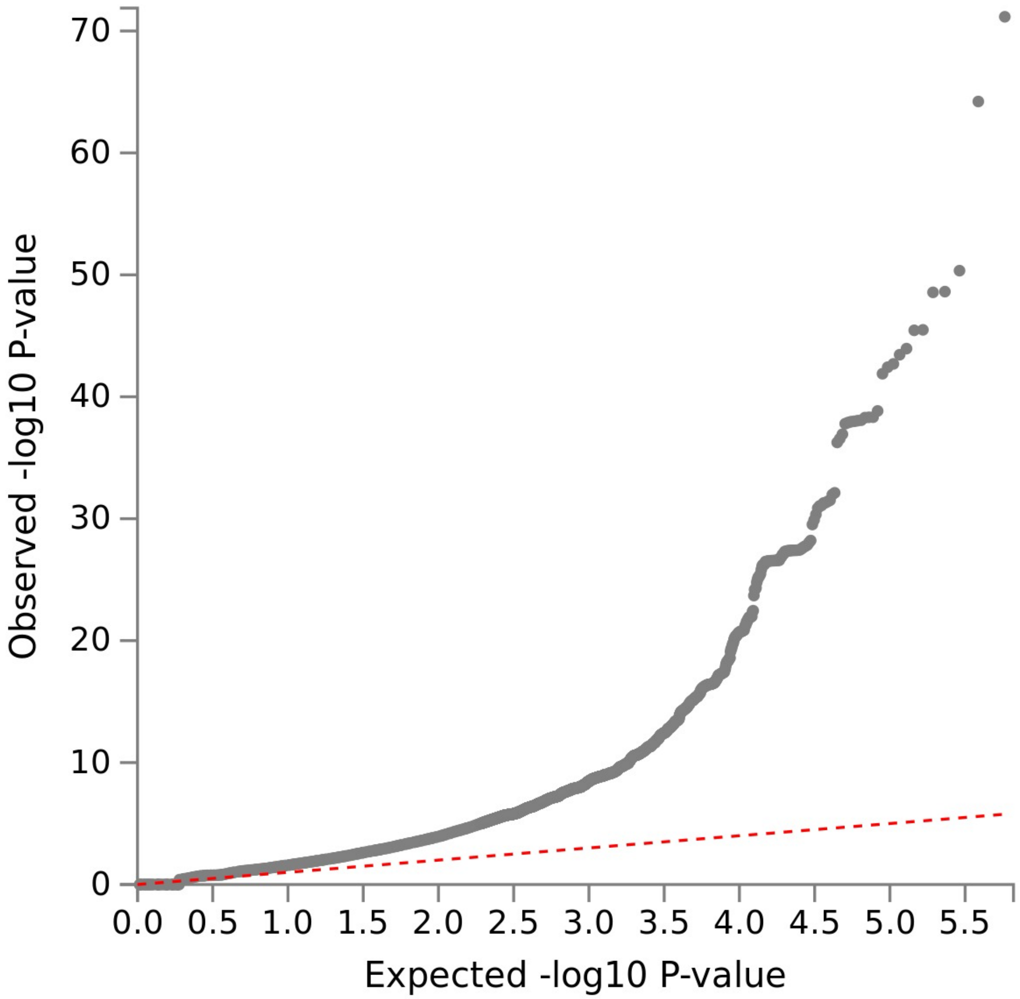


**Suppl. Figure 1.** Quantile-quantile (QQ) plot for the joint GWAS analysis shown in **Figure 1C**.

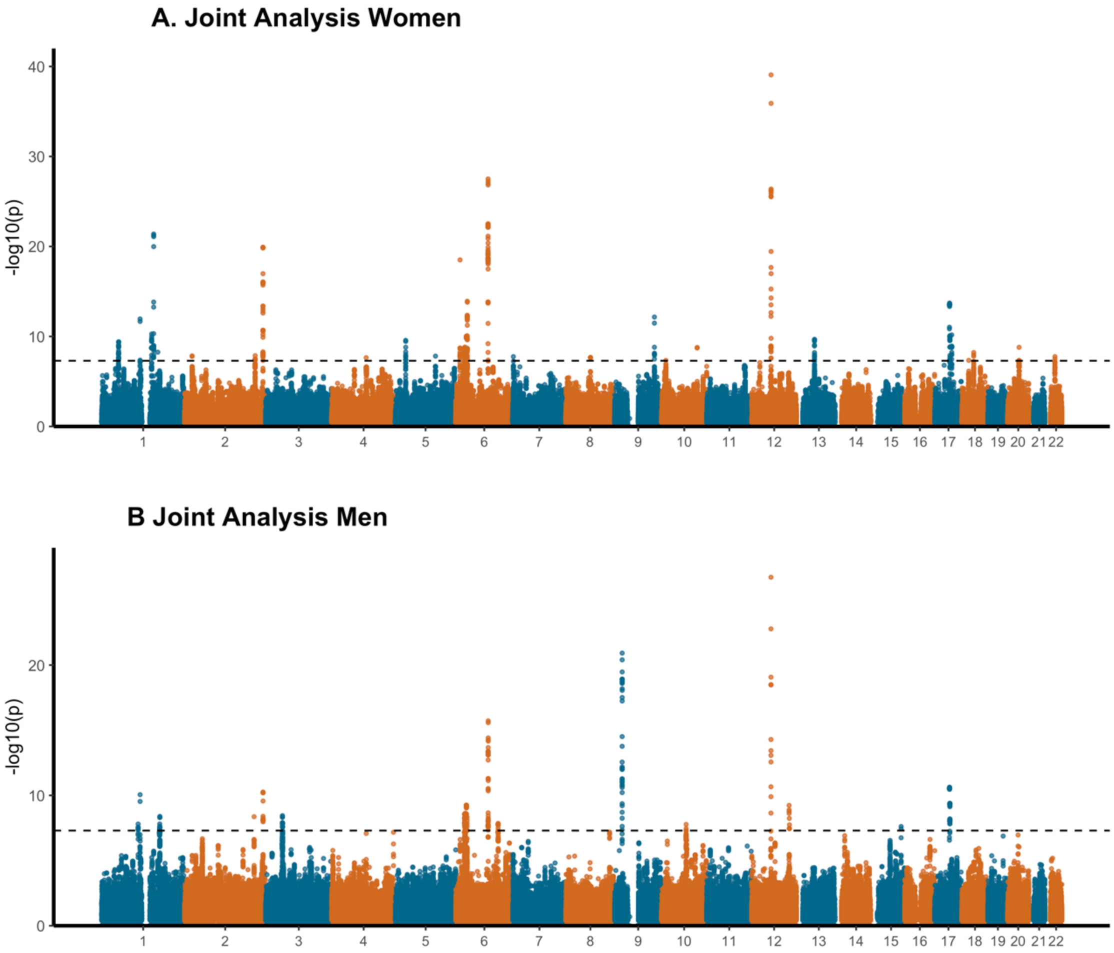


**Suppl. Figure 2. Sex-specific joint GWAS analysis of pain-related traits. A.** Manhattan plot of the meta-analysis performed in women only samples. **B.** Manhattan plot of the meta-analysis performed in men only samples.

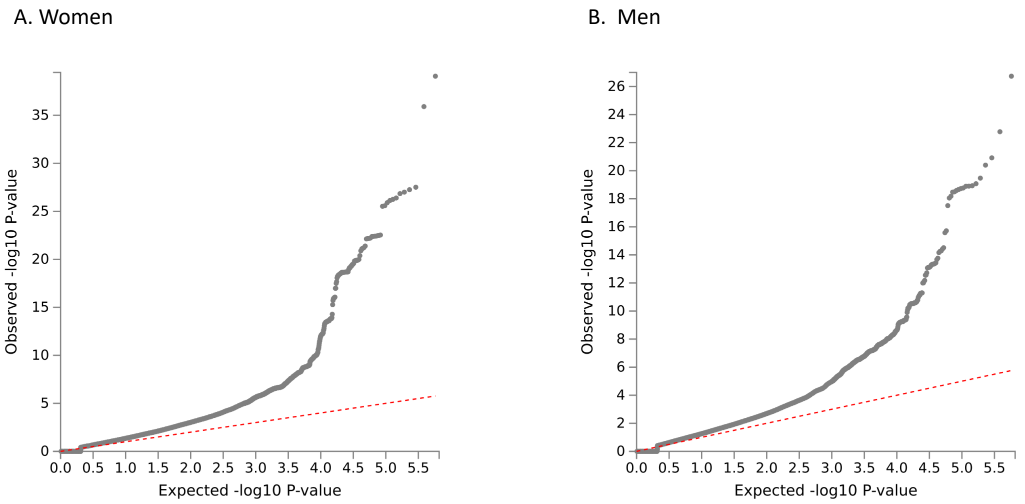


**Suppl. Figure 3. Quantile-quantile (QQ) plots for sex-specific analyses.** A. Female-only. B. Male-only.

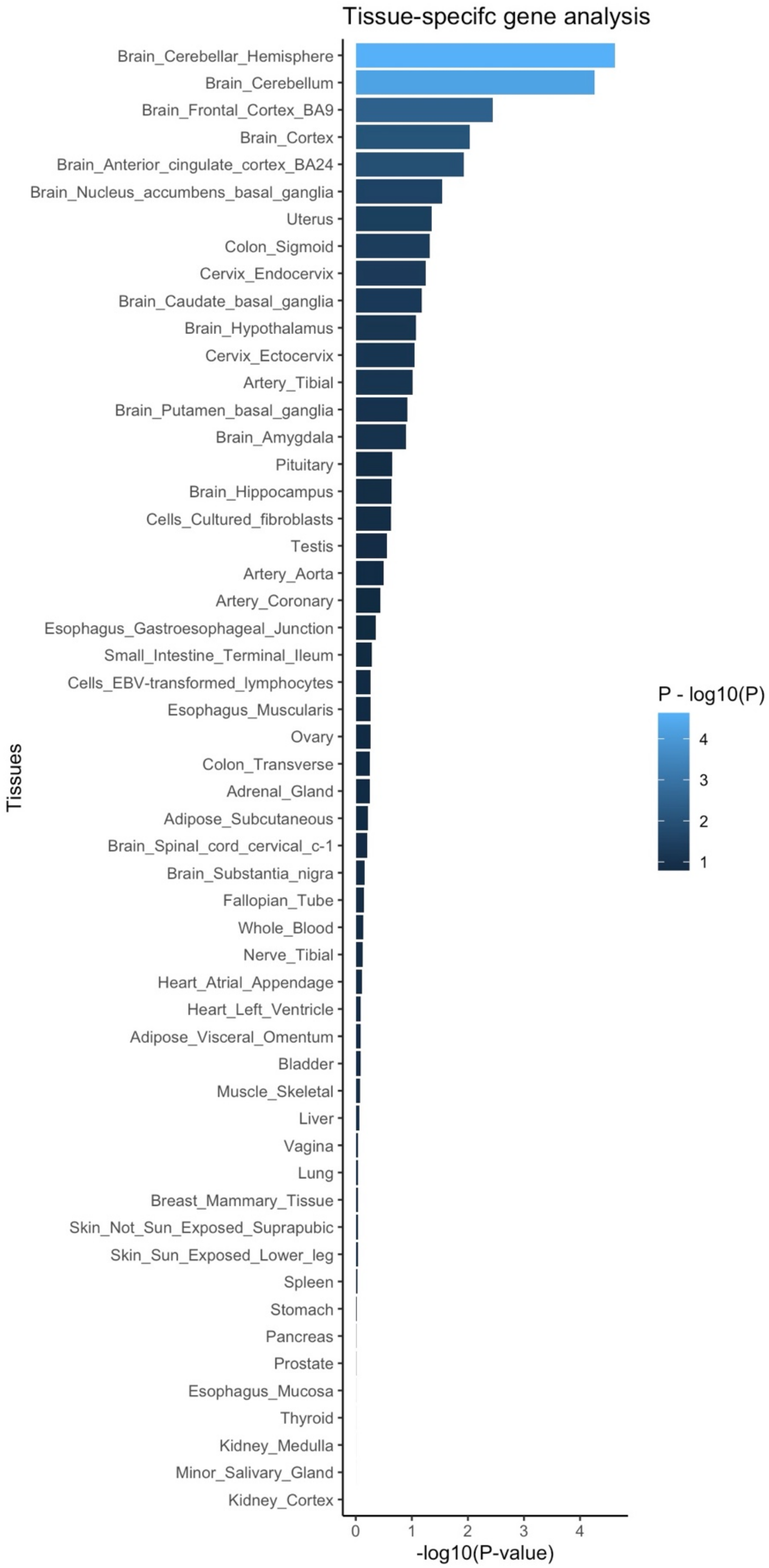


**Suppl. Figure 4. Enrichment of pain risk loci for tissue-specific gene expression.** The X axis indicates the significance of the test and in the Y axis indicates tissues analyzed.

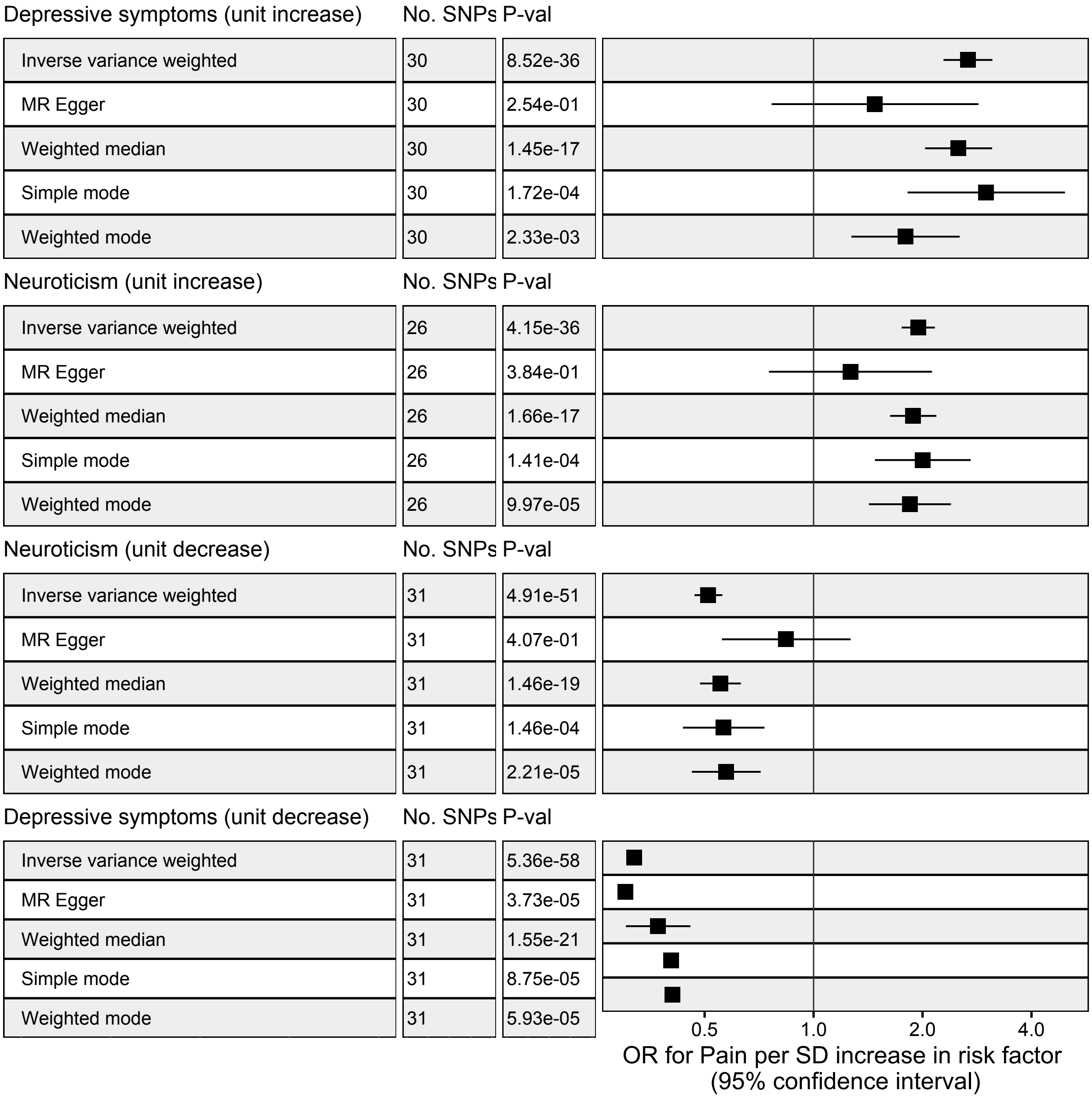


**Suppl. Figure 5. Two-sample Mendelian randomization results for psychiatric traits.** The X axis measures the odds ratio and its 95% confidence interval for pain per standard deviation (SD) increase or decrease of the risk factor.

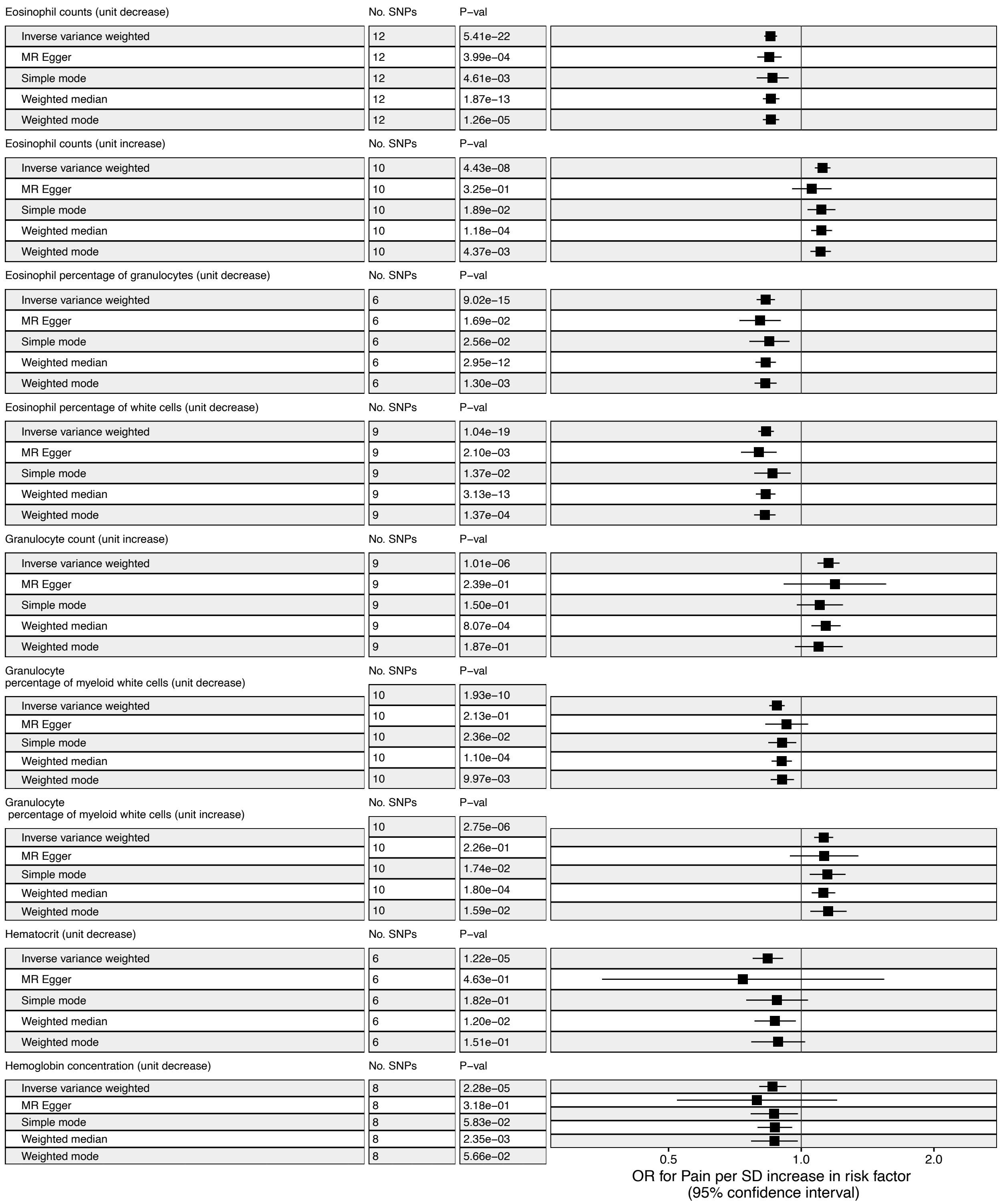


**Suppl. Figure 6. Two-sample Mendelian randomization results for immunological traits.** The X axis measures the odds ratio and its 95% confidence interval for pain per standard deviation (SD) increase or decrease of the risk factor. (continued next three pages)

| Hemoglobin concentration (unit increase) | No. SNPs | P−val |
|---|---|---|
| Inverse variance weighted | 6 | 9.79e−06 |
| MR Egger | 6 | 7.58e−01 |
| Simple mode | 6 | 7.09e−02 |
| Weighted median | 6 | 1.77e−04 |
| Weighted mode | 6 | 2.19e−02 |

| Immature fraction of reticulocytes (unit decrease) | No. SNPs | P−val |
|---|---|---|
| Inverse variance weighted | 9 | 5.74e−08 |
| MR Egger | 9 | 8.10e−02 |
| Simple mode | 9 | 6.76e−02 |
| Weighted median | 9 | 3.13e−04 |
| Weighted mode | 9 | 7.83e−02 |

| Immature fraction of reticulocytes (unit increase) | No. SNPs | P−val |
|---|---|---|
| Inverse variance weighted | 4 | 4.56e−04 |
| MR Egger | 4 | 3.76e−01 |
| Simple mode | 4 | 1.53e−01 |
| Weighted median | 4 | 2.92e−03 |
| Weighted mode | 4 | 6.51e−02 |

| Lymphocyte counts (unit increase) | No. SNPs | P−val |
|---|---|---|
| Inverse variance weighted | 9 | 1.35e−09 |
| MR Egger | 9 | 1.57e−01 |
| Simple mode | 9 | 1.23e−02 |
| Weighted median | 9 | 4.38e−05 |
| Weighted mode | 9 | 8.21e−03 |

| Mean corpuscular hemoglobin concentration (unit decrease) | No. SNPs | P−val |
|---|---|---|
| Inverse variance weighted | 4 | 2.18e−03 |
| MR Egger | 4 | 6.82e−01 |
| Simple mode | 4 | 9.77e−02 |
| Weighted median | 4 | 7.68e−03 |
| Weighted mode | 4 | 1.10e−01 |

| Mean corpuscular volume (unit increase) | No. SNPs | P−val |
|---|---|---|
| Inverse variance weighted | 13 | 1.91e−10 |
| MR Egger | 13 | 2.34e−01 |
| Simple mode | 13 | 1.54e−02 |
| Weighted median | 13 | 2.12e−05 |
| Weighted mode | 13 | 8.80e−03 |

| Mean platelet volume (unit increase) | No. SNPs | P−val |
|---|---|---|
| Inverse variance weighted | 12 | 8.72e−10 |
| MR Egger | 12 | 2.02e−01 |
| Simple mode | 12 | 7.57e−02 |
| Weighted median | 12 | 2.32e−03 |
| Weighted mode | 12 | 9.82e−02 |

| Monocyte percentage of white cells (unit decrease) | No. SNPs | P−val |
|---|---|---|
| Inverse variance weighted | 7 | 6.29e−05 |
| MR Egger | 7 | 5.30e−01 |
| Simple mode | 7 | 5.51e−02 |
| Weighted median | 7 | 1.40e−03 |
| Weighted mode | 7 | 5.37e−02 |

| Monocyte percentage of white cells (unit increase) | No. SNPs | P−val |
|---|---|---|
| Inverse variance weighted | 9 | 2.05e−08 |
| MR Egger | 9 | 2.04e−01 |
| Simple mode | 9 | 4.57e−02 |
| Weighted median | 9 | 6.82e−05 |
| Weighted mode | 9 | 2.73e−02 |

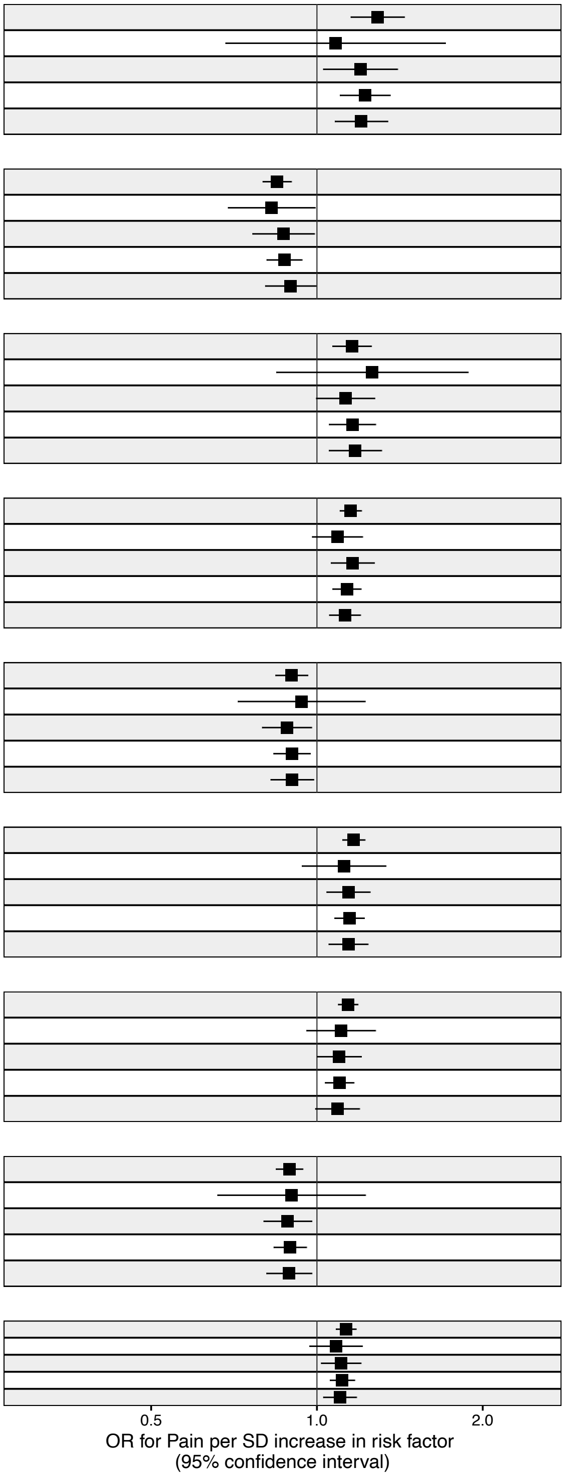


**Suppl. Figure 6. Two-sample Mendelian randomization results for immunological traits.** The X axis measures the odds ratio and its 95% confidence interval for pain per standard deviation (SD) increase or decrease of the risk factor. (page 2)

| Myeloid white cell count (unit increase) | No. SNPs | P–val |
|---|---|---|
| Inverse variance weighted | 8 | 3.21e–06 |
| MR Egger | 8 | 6.13e–01 |
| Simple mode | 8 | 2.57e–02 |
| Weighted median | 8 | 1.96e–04 |
| Weighted mode | 8 | 3.47e–01 |

| Neutrophil count (unit increase) | No. SNPs | P–val |
|---|---|---|
| Inverse variance weighted | 7 | 4.85e–06 |
| MR Egger | 7 | 7.85e–01 |
| Simple mode | 7 | 5.25e–02 |
| Weighted median | 7 | 6.34e–05 |
| Weighted mode | 7 | 4.75e–02 |

| Neutrophil percentage of granulocytes (unit decrease) | No. SNPs | P–val |
|---|---|---|
| Inverse variance weighted | 10 | 4.37e–08 |
| MR Egger | 10 | 1.77e–01 |
| Simple mode | 10 | 1.92e–02 |
| Weighted median | 10 | 4.75e–05 |
| Weighted mode | 10 | 7.06e–03 |

| Neutrophil percentage of granulocytes (unit increase) | No. SNPs | P–val |
|---|---|---|
| Inverse variance weighted | 4 | 6.61e–09 |
| MR Egger | 4 | 1.42e–01 |
| Simple mode | 4 | 3.67e–02 |
| Weighted median | 4 | 1.20e–11 |
| Weighted mode | 4 | 6.78e–03 |

| Neutrophil percentage of white cells (unit decrease) | No. SNPs | P–val |
|---|---|---|
| Inverse variance weighted | 7 | 5.41e–06 |
| MR Egger | 7 | 2.75e–01 |
| Simple mode | 7 | 1.52e–02 |
| Weighted median | 7 | 1.50e–04 |
| Weighted mode | 7 | 1.96e–02 |

| Neutrophil percentage of white cells (unit increase) | No. SNPs | P–val |
|---|---|---|
| Inverse variance weighted | 7 | 7.97e–05 |
| MR Egger | 7 | 4.56e–01 |
| Simple mode | 7 | 1.25e–01 |
| Weighted median | 7 | 9.82e–03 |
| Weighted mode | 7 | 9.20e–02 |

| Plateletcrit (unit increase) | No. SNPs | P–val |
|---|---|---|
| Inverse variance weighted | 7 | 5.73e–07 |
| MR Egger | 7 | 4.79e–01 |
| Simple mode | 7 | 9.10e–02 |
| Weighted median | 7 | 7.14e–04 |
| Weighted mode | 7 | 7.56e–02 |

| Red cell distribution width (unit decrease) | No. SNPs | P–val |
|---|---|---|
| Inverse variance weighted | 7 | 3.46e–05 |
| MR Egger | 7 | 3.40e–01 |
| Simple mode | 7 | 7.38e–02 |
| Weighted median | 7 | 6.92e–04 |
| Weighted mode | 7 | 5.22e–02 |

| Red cell distribution width (unit increase) | No. SNPs | P–val |
|---|---|---|
| Inverse variance weighted | 8 | 4.98e–04 |
| MR Egger | 8 | 2.74e–01 |
| Simple mode | 8 | 8.03e–02 |
| Weighted median | 8 | 8.55e–03 |
| Weighted mode | 8 | 9.28e–02 |

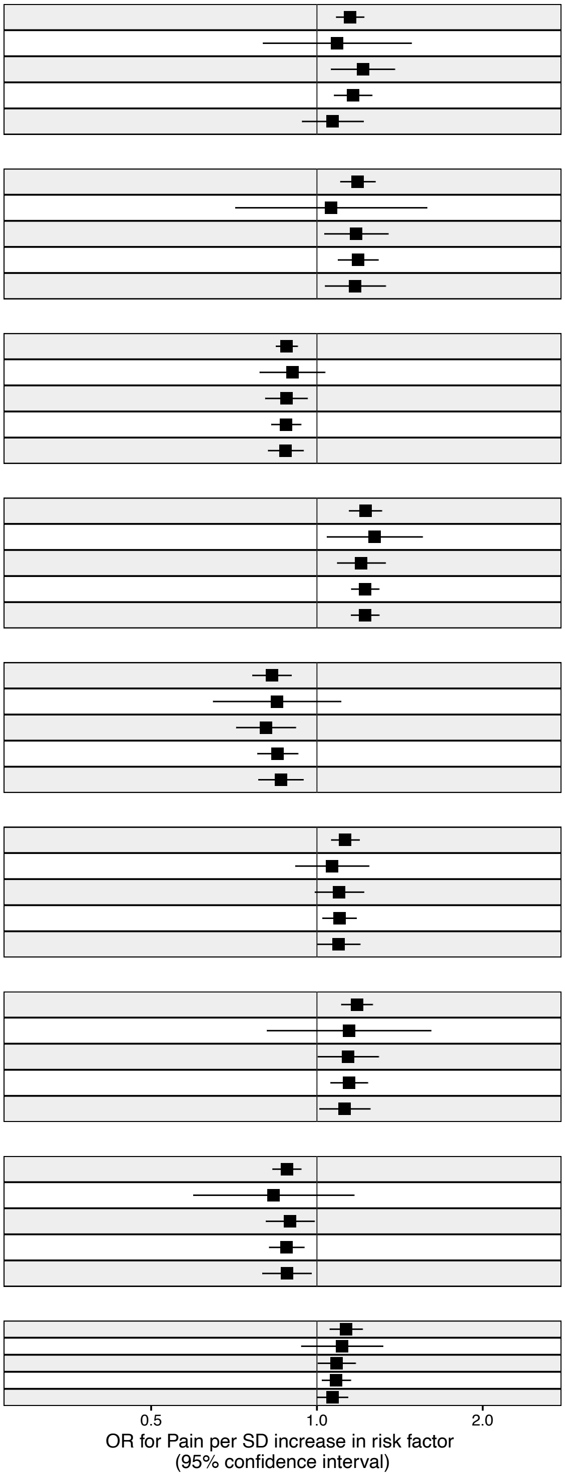


**Suppl. Figure 6. Two-sample Mendelian randomization results for immunological traits.** The X axis measures the odds ratio and its 95% confidence interval for pain per standard deviation (SD) increase or decrease of the risk factor. (page 3)

Reticulocyte fraction of red cells (unit decrease)

| | No. SNPs | P−val |
|---|---|---|
| Inverse variance weighted | 10 | 1.85e−04 |
| MR Egger | 10 | 4.72e−01 |
| Simple mode | 10 | 1.24e−02 |
| Weighted median | 10 | 1.07e−02 |
| Weighted mode | 10 | 2.01e−02 |

Sum basophil neutrophil counts (unit increase)

| | No. SNPs | P−val |
|---|---|---|
| Inverse variance weighted | 8 | 2.41e−06 |
| MR Egger | 8 | 6.46e−01 |
| Simple mode | 8 | 6.66e−02 |
| Weighted median | 8 | 4.67e−04 |
| Weighted mode | 8 | 6.97e−02 |

Sum eosinophil basophil counts (unit decrease)

| | No. SNPs | P−val |
|---|---|---|
| Inverse variance weighted | 10 | 1.42e−17 |
| MR Egger | 10 | 5.87e−03 |
| Simple mode | 10 | 1.89e−03 |
| Weighted median | 10 | 6.32e−15 |
| Weighted mode | 10 | 2.20e−05 |

Sum neutrophil eosinophil counts (unit increase)

| | No. SNPs | P−val |
|---|---|---|
| Inverse variance weighted | 8 | 5.10e−06 |
| MR Egger | 8 | 1.73e−01 |
| Simple mode | 8 | 1.42e−01 |
| Weighted median | 8 | 5.43e−03 |
| Weighted mode | 8 | 2.00e−01 |

White blood cell count (unit increase)

| | No. SNPs | P−val |
|---|---|---|
| Inverse variance weighted | 9 | 3.53e−07 |
| MR Egger | 9 | 2.52e−01 |
| Simple mode | 9 | 1.13e−01 |
| Weighted median | 9 | 1.88e−04 |
| Weighted mode | 9 | 2.61e−02 |

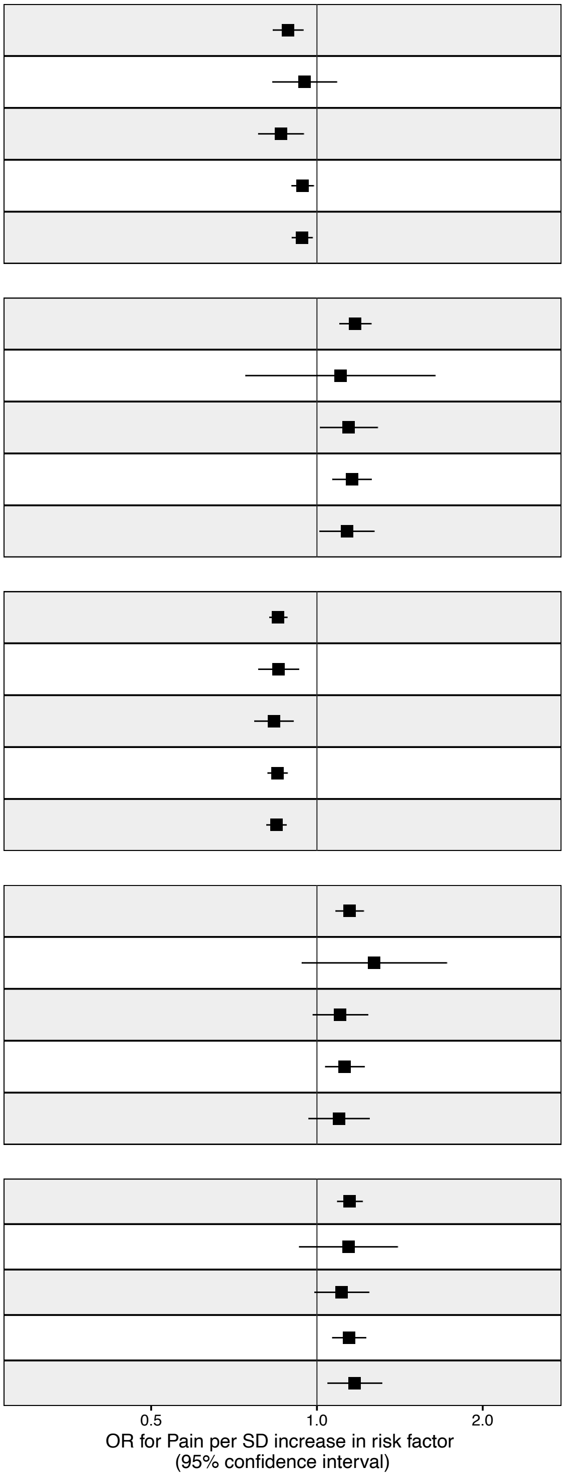


**Suppl. Figure 6. Two-sample Mendelian randomization results for immunological traits.** The X axis measures the odds ratio and its 95% confidence interval for pain per standard deviation (SD) increase or decrease of the risk factor. (page 4)

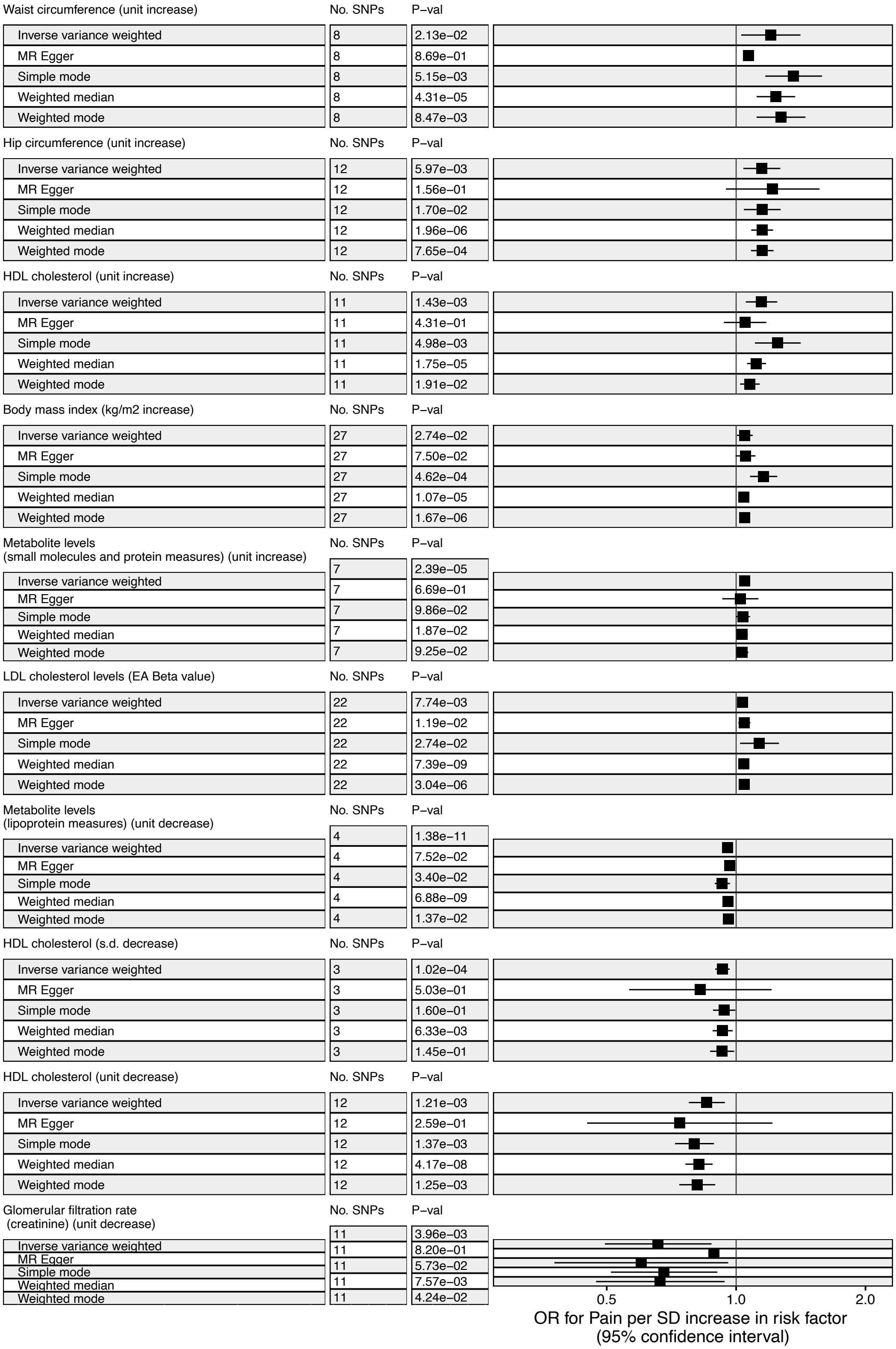


**Suppl.Figure 7. Two-sample Mendelian randomization results for metabolic traits.** The X axis measures the odds ratio and its 95% confidence interval for pain per standard deviation (SD) increase or decrease of the risk factor.

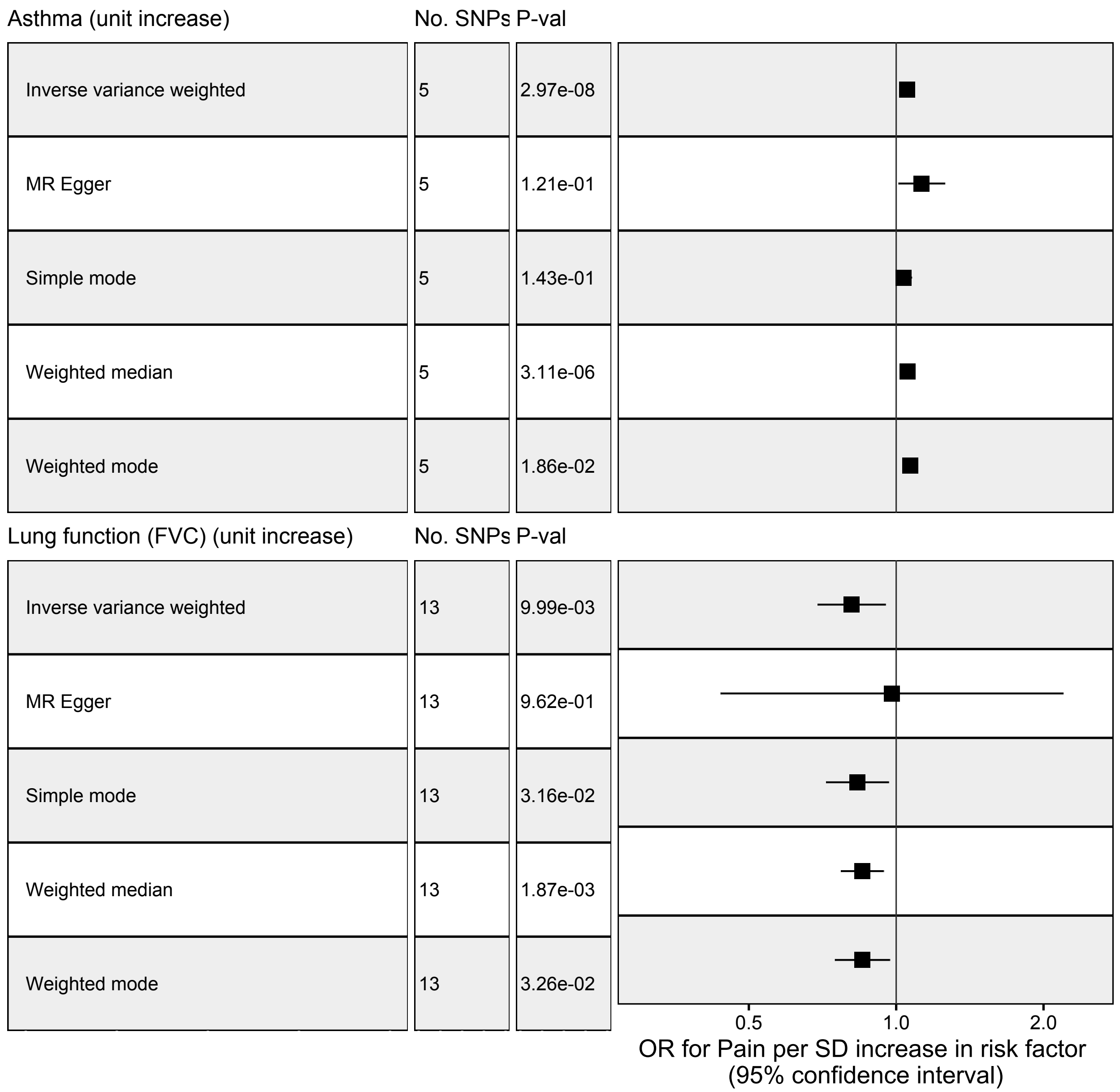


**Suppl. Figure 8. Two-sample Mendelian randomization results for respiratory traits.** The X axis measures the odds ratio and its 95% confidence interval for pain per standard deviation (SD) increase or decrease of the risk factor.

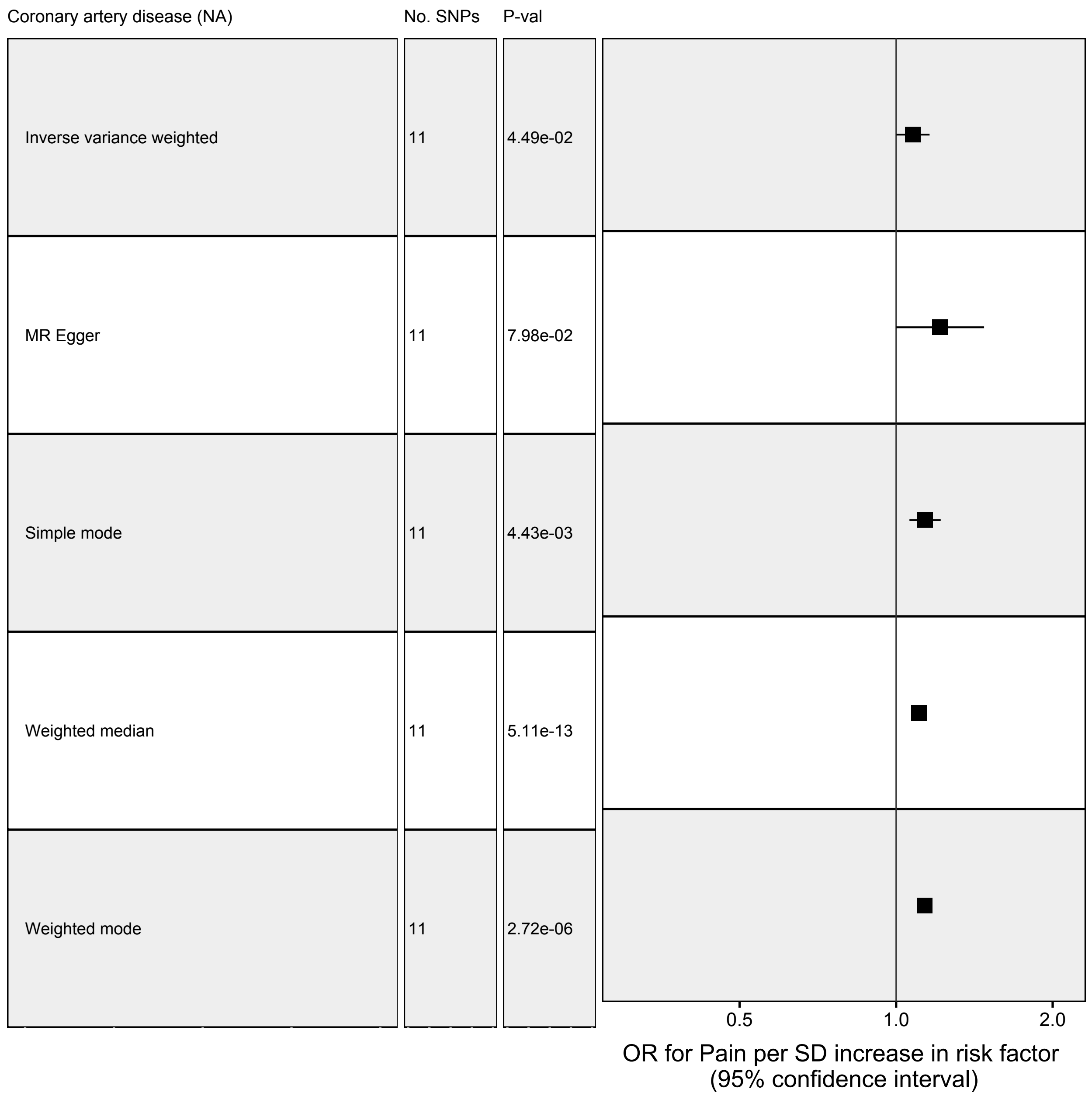


**Suppl. Figure 9. Two-sample Mendelian randomization results for cardiovascular traits.** The X axis measures the odds ratio and its 95% confidence interval for pain per standard deviation (SD) increase or decrease of the risk factor.

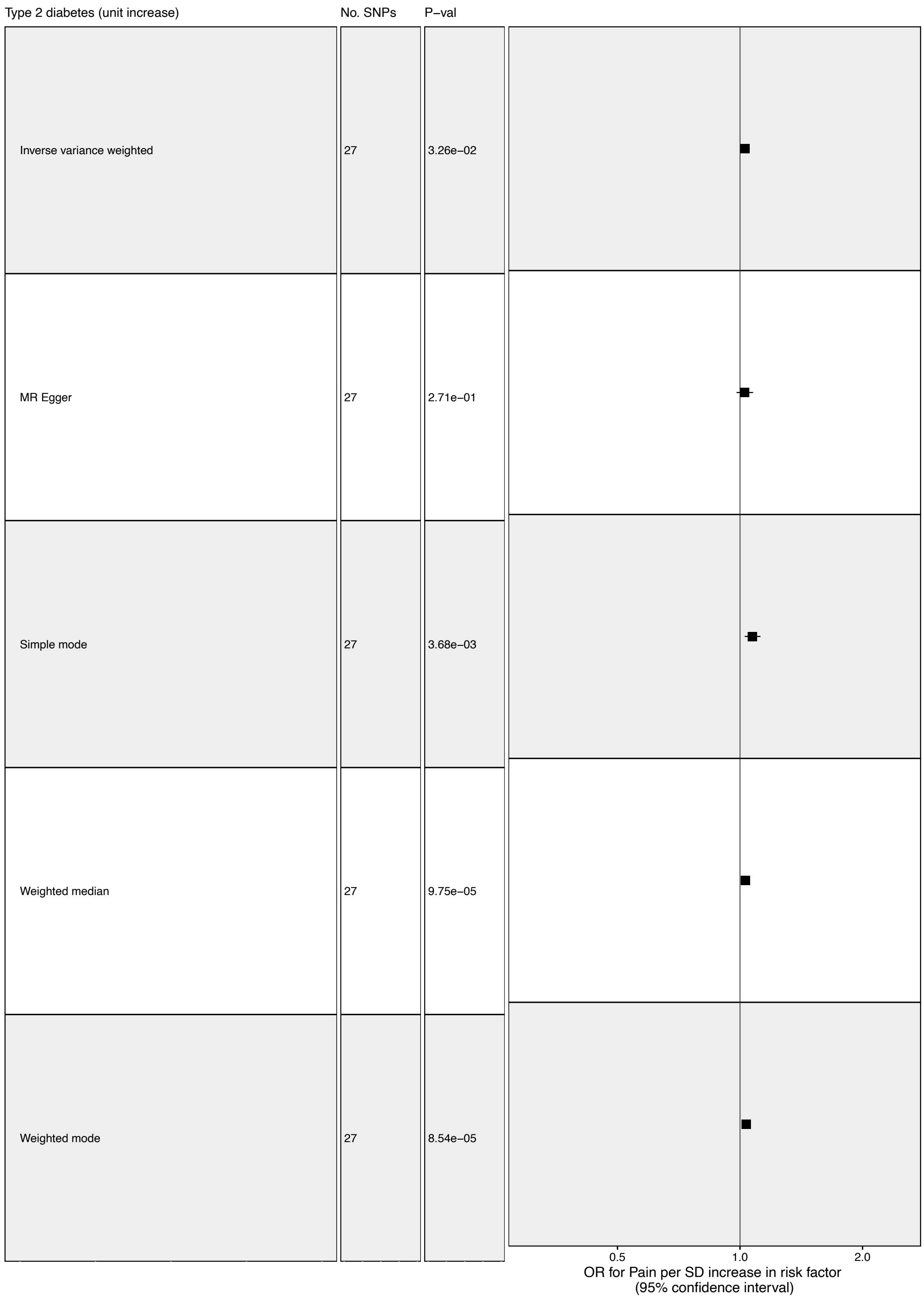


**Suppl. Figure 10. Two-sample Mendelian randomization results for endocrine traits.** The X axis measures the odds ratio and its 95% confidence interval for pain per standard deviation (SD) increase or decrease of the risk factor.

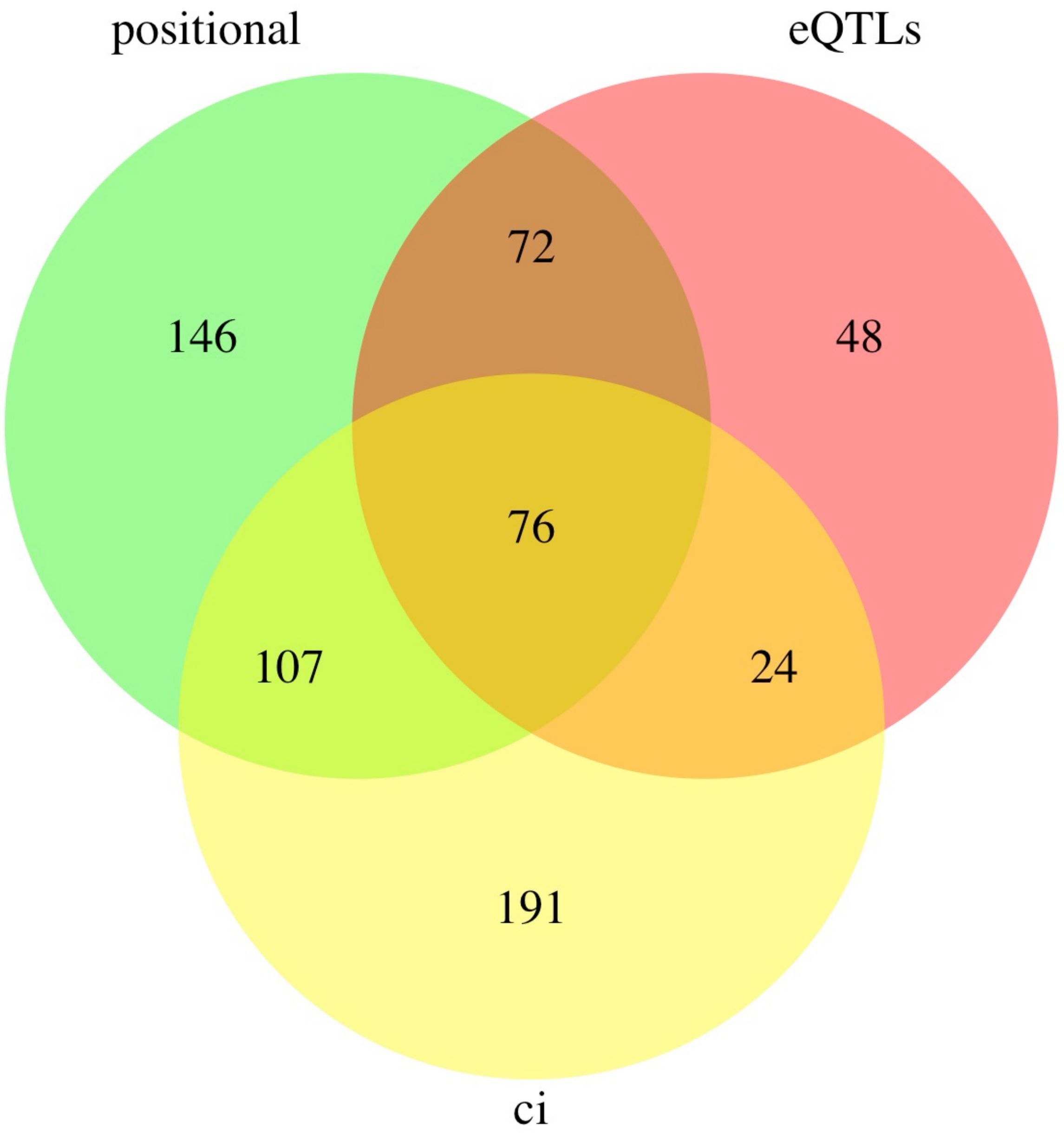


**Suppl. Figure 11.** Venn diagram showing the overlap of genes selected by position, eQTLs and chromatin interaction gene mapping methods

**Supplementary Tables**

**Supplementary Table 1. Pain phenotypes**. This supplementary table contains details of the ascertainment and GWAS analyses for 17 selected pain phenotypes. Ten pain phenotypes were created through a touchscreen questionnaire with the main question: "pain types experienced in the last month" (field ID 6159). The possible answers were: 'None of the above' (renamed in the present study as "pain any" and coded as 6159_100); 'Prefer not to answer' (6159_9); pain at seven different body sites (head (6159_1)), face (6159_2), neck/shoulder (6159_3), back (6159_4), stomach/abdominal (6159_5), hip (6159_6), knee (6159_7 ); or 'all over the body' (6159_8) . With the exception of the field 6159_8, all the others are not mutually exclusive since a participant can experience pain in multiple parts of the body and thereby, they can select multiple answers. Patients that experienced pain in the last month were further asked whether the pain had lasted for 3 months or longer. This condition is described as chronic pain and indicated with the category ID 100048; for this study we selected back and headache chronic pain sites, coded as "3571" and "3799" respectively. In addition, we included five traits symbolized with code "6154", specify distinct pain relief medications: Aspirin (6154_1), Ibuprofen (6154_2), Paracetamol (6154_3), Omeprazol (6154_5) and in addition Medication none of the above (that we named as medication any) (6154_100). Finally, we included two pain-related phenotypes to our combined analysis, "chest pain or discomfort" marked with the code "2335" and extracted from the category chest pain and "leg pain on walking" derived from the category claudication and peripheral artery disease and identified with the code "4728".

**Supplementary Table 2. Heritability estimates for pain phenotypes.** Heritability estimates are reproduced from analyses by the Neale lab at Massachusetts General Hospital using LDSC. In this study we included only traits with statistically significant heritability estimates, z>4.

**Supplementary Table 3. Pairwise genetic correlation among pain phenotypes.** Pairwise genetic correlations among the 17 pain UKBB traits were calculated with LDSC. Values range from -1 to 1 with estimates close to -1 indicating high negative genetic correlation, while values close to 1 indicate strong positive genetic correlation between traits.

**Supplementary Table 4. Risk loci from joint analysis of pain-related traits.** Description of 99 genome-wide significant loci ($P$ < 5e-8) identified in the joint analysis of the 17 pain UKBB traits.

**Supplementary Table 5. Overlap of risk loci from joint analysis vs. underlying pain-related traits.** Number of genome-wide significant loci by trait. In green are indicated the loci that did not show any significant locus for underlying traits (N=51). In beige are displayed loci that reported one significant locus (N=37). Highlighted in lilac are traits with two genome-wide significant loci (N=4). Traits colored in brown included 3 significant loci (N=3). Brick and blue colors represent traits with 4 and 6 significant loci respectively (N=1 each).

**Supplementary Table 6. Novel pain genomic risk loci**. To the best of our knowledge the loci listed in this table have not been previously associated to pain phenotype.

**Supplementary Table 7. Overlap with previously reported risk loci.** Thirty-seven of the 99 loci were previously reported associated to pain phenotypes. Of them, 10 were found in Multisite chronic pain (pink), 20 in migraine (orange), 1 in Neck pain or shoulder pain (lilac), Chronic back pain (blue), and knee pain (yellow). Finally, 3 loci were previously found associated to both migraine and Multisite chronic pain (light green) and 1 in Multisite chronic pain & Neck pain or shoulder pain (dark green).

**Supplementary Table 8. Pseudo-replication in male vs. female samples.** The 99 genome-wide significant loci leading, and highly related SNPs were checked for replication (P<=0.05) separately in men and women samples.

**Supplementary Table 9. Gene-based GWAS p-values.** Whole genome gene-based analysis performed using MAGMA implemented in FUMA. Genes were considered statistically significant if their association P-value was < 2.8x10-6. The table lists genes in order of decreasing significance.

**Supplementary Table 10.** This table lists 46 genes that showed genome-wide significant at the gene-based analysis and that were also the closest genes to the leading and/or their highly related SNPs from the 99 genome-wide significant loci.

**Supplementary Table 11. Public databases of pain genes. Sheet 1.** Human pain genes. The table shows a comprehensive description of the genes associated with pain in human studies, including clinical studies, candidate gene and GWAS analyses; overall, up to 2018, 353 genes have been included in this list (Meloto et al., 2018). **Sheet 2.** Mouse pain genes. This table includes a list of genes identified associated with pain through mouse experiments. Gene collection has been discontinued in 2015 (Lacroix-Fralish ML et al., 2007) and the database includes 434 genes.

**Supplementary Table 12. G**ene set enrichment analysis results Gene Ontology Biological Process, Molecular Function, and Cellular Component terms, and for KEGG and Reactome.

**Supplementary Table 13. Annotation of neuronal genes at pain risk loci.** Sheets indicate the genes from GO terms neurogenesis (A), neurodifferentiation (B), and post synapsis (C) with significant associations to pain. For each gene included in each gene set we show its gene-based p-value from MAGMA.

**Supplementary Table 14. Pleiotropic effects of pain risk SNPs on non-pain traits. Pleiotropic effects were mapped by integration with the GWAS Atlas. Traits are grouped in 21 categor**ies listed on different sheets in the table.

**Supplementary Table 15. Genetic correlations with non-pain traits. Genetic correlations were calculated between pain and traits in the LDHub database (Sheet A), as well as between** pain and several immunological traits for which we had detected pleiotropic effects at genome-wide significant risk loci (Sheet B)**.**

**Supplementary Table 16. Two-sample Mendelian randomization.** The table lists TwoSampleMR results obtained by analyzing several categories of non-pain traits as exposure (i.e., risk factor) with pain as the outcome. For each trait the table lists results for the primary inverse variance weighted (IVW) test, followed by the results obtained using the sensitivity tests: MR Egger, weighted median and simple and weighted mode. When the results suggest pleiotropy, both IVW and MR Egger are tested for heterogeneity among their single SNPs estimates using Cochran Q heterogeneity test. Pleiotropy is measured as the intercept term in the MR Egger method and interpreted as an estimate of the average pleiotropic effect across all genetic variants. **A.** TwoSampleMR using psychiatric traits as exposure and pain as outcome **B.** TwoSampleMR using immunological traits as exposure and pain as outcome. **C.** TwoSampleMR using metabolites traits as exposure and pain as outcome. **D.** TwoSampleMR using respiratory traits as exposure and pain as outcome. **E.** TwoSampleMR using cardiovascular traits as exposure and pain as outcome**. F.** TwoSampleMR using endocrine traits as exposure and pain as outcome

**Supplementary Table 17. Genes at risk loci prioritized by proximity, eQTLs, and chromatin interactions. A.** Positional gene mapping. B. Expression qiantitative trait loci (eQTLs) gene mapping**. C.** Chromatin interaction gene mapping**. D.** Intersection of genes identified through positional and eQTLs gene mapping**. E. Intersection of** genes identified through eQTLs and chromatin interaction gene mapping**. F. Intersection of** genes identified through positional and chromatin interaction gene mapping**. G. Intersection of g**enes identified through all three gene mapping methods

**Supplementary Table 18. Single SNP TwoSampleMR analysis.** Note: we considered results to be significant only when the SNPs reached genome-wide significance for association both with pain and with the predicted mediating factor.
